# Full-spectrum dynamics of the coronavirus disease outbreak in Wuhan, China: a modeling study of 32,583 laboratory-confirmed cases

**DOI:** 10.1101/2020.04.27.20078436

**Authors:** Xingjie Hao, Shanshan Cheng, Degang Wu, Tangchun Wu, Xihong Lin, Chaolong Wang

## Abstract

Vigorous non-pharmaceutical interventions have largely suppressed the COVID-19 outbreak in Wuhan, China. We extended the susceptible-exposed-infectious-recovered model to study the transmission dynamics and evaluate the impact of interventions using 32,583 laboratory-confirmed cases from December 8, 2019 till March 8, 2020, accounting for presymptomatic infectiousness, and time-varying ascertainment rates, transmission rates, and population movements. The effective reproduction number R_0_ dropped from 3.54 (95% credible interval: 3.41-3.66) in the early outbreak to 0.27 (0.23-0.32) after full-scale multi-pronged interventions. By projection, the interventions reduced the total infections in Wuhan by 96.1% till March 8. Furthermore, we estimated that 87% infections (lower bound: 53%) were unascertained, potentially including asymptomatic and mild-symptomatic cases. The probability of resurgence was 0.33 and 0.06 based on models with 87% and 53% infections unascertained, respectively, assuming all interventions were lifted after 14 days of no ascertained infections. These results provide important implications for continuing surveillance and interventions to eventually contain the outbreak.

---

The coronavirus disease 2019 (COVID-19) caused by SARS-CoV-2 was detected in Wuhan, China, in December 2019.^1^ Many early cases were connected to the Huanan Seafood Market, which was disinfected on January 1, 2020 to stop potential zoonotic infection.^2^ Nevertheless, the high population density of Wuhan together with the increased social activities before the Chinese New Year catalyzed the outbreak in January, 2020. The massive human movement during the holiday travel season *Chunyun*, which started on January 10, further expedited spreading of the outbreak.^3^ Shortly after the confirmation of human-to-human transmission, the Chinese authorities implemented the unprecedented *cordons sanitaire* of Wuhan on January 23 to contain the geographic spread, followed by a series of non-pharmaceutical interventions to reduce virus transmission, including suspension of all intra- and inter-city transportation, compulsory mask wearing in public places, cancelation of social gatherings, and home quarantine of mild-symptomatic patients.^4^ From February 2, strict stay-at-home policy for all residents, centralized isolation of all patients, and centralized quarantine of suspected cases and close contacts were implemented to stop household and community transmission. Furthermore, a city-wide door-to-door universal symptom survey was carried out during February 17-19 by designated community workers to identify previously undetected symptomatic cases. Details of the interventions were described in Pan *et al*.^4^ These drastic interventions, together with the improved medical resources and healthcare manpower from all over the country, have effectively bent the epidemic curve and reduced the attack rate in Wuhan, shedding light on the global efforts to control the COVID-19 outbreak.^4^

Recent studies have revealed important transmission features of COVID-19, including infectiousness of asymptomatic cases^5-9^ and presymptomatic cases.^10-12^ Furthermore, the number of ascertained cases was much smaller than those estimated by earlier modeling-based studies using international cases exported from Wuhan prior to the travel suspension,^3,13,14^ implying a substantial number of unascertained cases. Using reported cases from 375 cities in China, a modeling study concluded that substantial unascertained cases, despite having lower transmissibility, had facilitated the rapid spreading of COVID-19.^15^ In addition, accounting for the unascertained cases has refined the estimation of case fatality risk of COVID-19, leading to a better understanding of the clinical severity of the disease.^16^ Modeling both ascertained and unascertained cases is important to facilitate interpretation of transmission dynamics and epidemic trajectories.

Based on comprehensive epidemiological data from Wuhan,^4^ we extended the susceptible-exposed-infectious-recovered (SEIR) model to delineate the full spectrum of COVID-19 outbreak in the epicenter, accounting for presymptomatic infectiousness, unascertained cases, population movement, and different intervention strengths across time periods (**Fig. 1)**. We named the extended model SAPHIRE, because we compartmentalized the population into *S* susceptible, *E* exposed, *P* presymptomatic, *A* unascertained, / ascertained, *H* isolated, and *R* removed individuals. Compared with the classic SEIR model, we explicitly modeled population movement^3^ and split the infectious cases into *P, A*, and *I* to reflect infectiousness at different stages. The unascertained compartment *A* was expected to mostly consist of asymptomatic and mild-symptomatic cases who were infectious but difficult to detect. We introduced compartment *H* because ascertained cases would have shorter effective infectious period due to isolation in the hospital, especially when medical resources were improved.

**Fig. 1.**
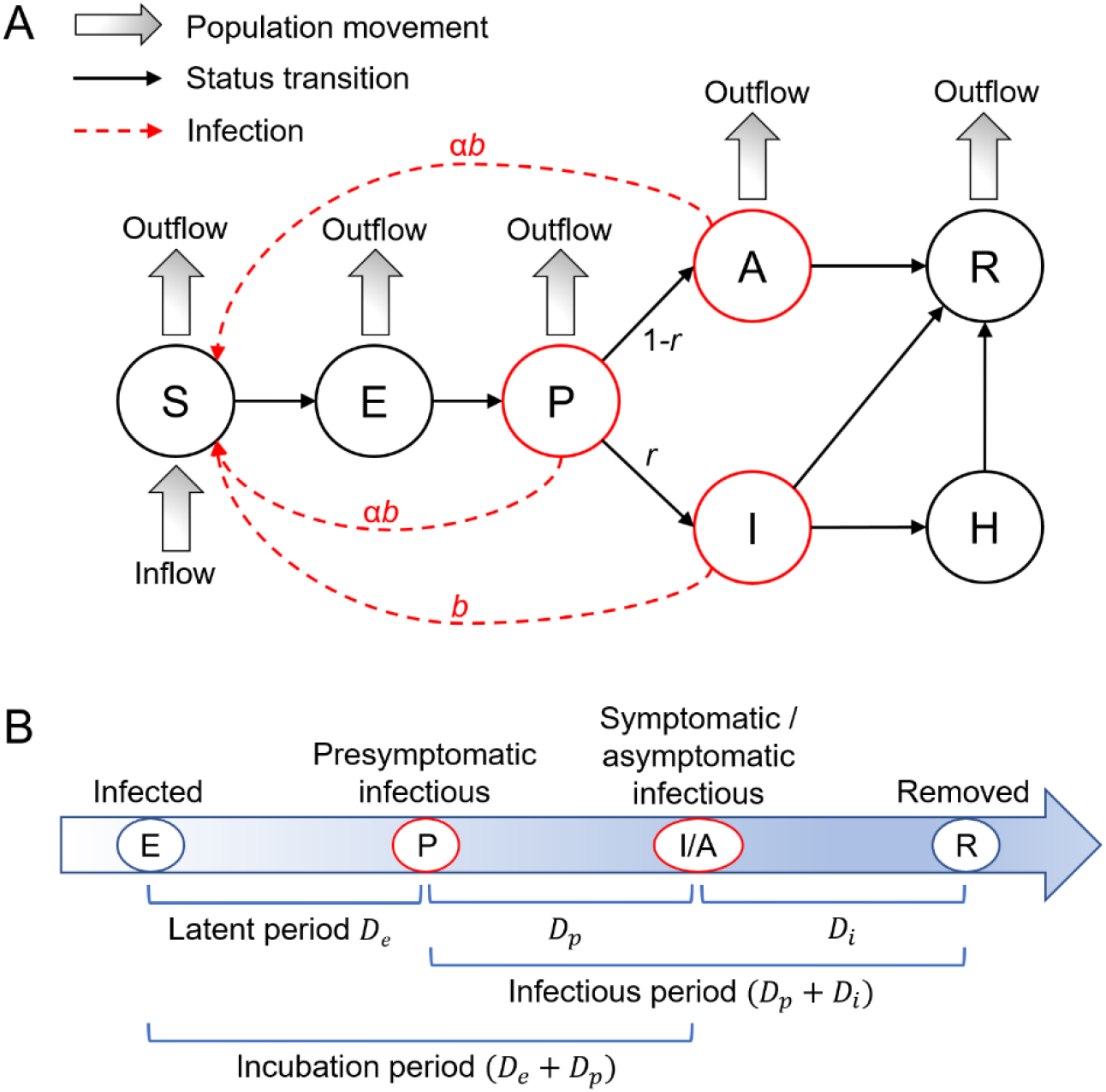
Illustration of the SAPHIRE model. We extended the classic SEIR model to include seven compartments, namely S (susceptible), E (exposed), P (presymptomatic infectious), I (ascertained infectious), A (unascertained infectious), H (isolated), and R (removed). (A) Relationship between different compartments in the model. Two parameters of interests are *r* (ascertainment rate) and *b* (transmission rate), which are assumed to be varying across time periods. (B) Schematic timeline of an individual from being exposed to the virus to recovery without isolation. In this model, the unascertained compartment A includes asymptomatic and some mild-symptomatic cases who were not detected. While there is no presymptomatic phase for asymptomatic cases, we treated asymptomatic as a special case of mild-symptomatic and modeled both with a “presymptomatic” phase for simplicity.

We chose to model the outbreak from January 1, 2020 and divided it into five time periods based on key events and interventions: January 1 to 9 (before *Chunyun*), January 10 to 22 (*Chunyun*), January 23 to February 1 (*cordons sanitaire*), February 2 to 16 (centralized isolation and quarantine), and February 17 to March 8 (community screening). We assumed a constant population size of 10 million with equal numbers of daily inbound and outbound travelers (500,000 before *Chunyun*, 800,000 during *Chunyun*, and 0 after *cordons sanitaire*).^3^ Furthermore, we assumed transmission rate and ascertainment rate did not change in the first two periods, because few interventions were implemented before January 23, while they were allowed to vary in later periods to reflect different intervention strengths. We used Markov Chain Monte Carlo (MCMC) to estimate these parameters by assuming the daily incidence following a Poisson distribution, while the other parameters were set based on previous epidemiological investigations^2,10^ or from our data (**Methods**). We assumed the transmissibility of presymptomatic/unascertained cases to be 0.55 of the ascertained cases.^15,17^

We first simulated epidemic curves with two periods to test the performance of our parameter estimation procedure (**Methods**). We converted the transmission rate to the effective reproduction number *R*_0_ and focused on evaluating the estimation of *R*_0_ and the ascertainment rate *r* in both periods, for which the parameter values were different. As shown in **Extended Data Figs. 1-2**, our method could accurately estimate *R*_0_ and the ascertainment rates when the model was correctly specified and was robust to misspecification of the duration from symptom onset to isolation and the relative transmissibility of presymptomatic/unascertained cases to ascertained cases. As expected, estimates of *R*_0_ were positively correlated with the specified latent period and infectious periods, while the estimated ascertainment rates were positively correlated with the specified ascertainment rate in the initial state (**Extended Data Fig. 2**). These simulation results highlighted the importance of carefully specifying parameter values and designing sensitivity analyses based on information from existing data and literature.

Based on confirmed cases exported from Wuhan to Singapore, we conservatively estimated the ascertainment rate during the early outbreak in Wuhan was 0.23 (95% confidence interval [CI]: 0.14-0.42) (**Methods**). We then applied our model to fit the daily incidences in Wuhan from January 1 to February 29, assuming the initial ascertainment rate was 0.23, and used the fitted model to predict the trend from March 1 to 8. As shown in **Fig. 2A**, our model fit the observed data well, except for the outlier on February 1, which might be due to approximate-date records of many patients admitted to the field hospitals set up after February 1. After a series of multi-faceted public health interventions, the transmission rate decreased from 1.31 (95% credible interval [CrI]: 1.25-1.37) in the first two periods to 0.40 (0.38-0.42), 0.17 (0.16-0.19), and 0.10 (0.08-0.12) in the later three periods, respectively (**Extended Data Table 5**), which could be translated into *R*_0_ of 3.54 (3.41-3.66), 3.32 (3.20-3.44), 1.18 (1.11-1.25), 0.51 (0.47-0.54) and 0.27 (0.23-0.32) for the five periods, respectively (**Fig. 2B, Extended Data Table 6**). We estimated the cumulative number of infections, including unascertained cases, till March 8 to be 257,406 (207,683-315,785) if the trend of the fourth period was assumed (**Fig. 2C**), or 812,930 (599,992-1,087,944) if the trend of the third period was assumed (**Fig. 2D**), or 6,302,928 (6,277,193-6,327,431) if the trend of the second period was assumed (**Fig. 2E**), in comparison to the estimated total infections of 248,022 (199,474-302,464) by fitting data from all five periods (**Fig. 2A**). These numbers were translated to 3.6%, 69.5%, and 96.1% reduction of infections due to the interventions in different periods.

**Fig. 2.**
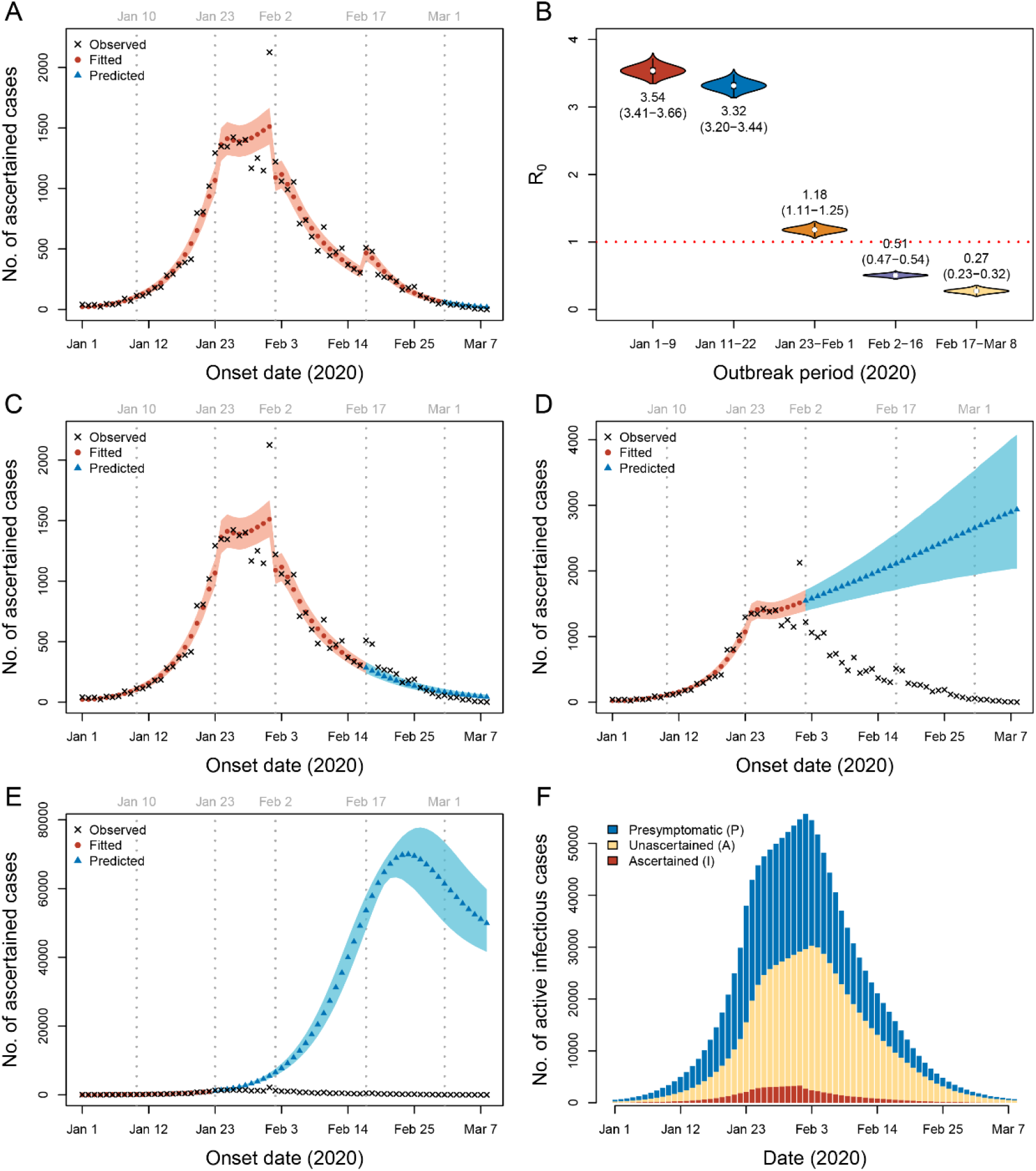
Modeling the COVID-19 epidemic in Wuhan. Parameters were estimated by fitting data from January 1 to February 29. (A) Prediction using parameters from period 5 (February 17-29). (B) Estimated R_0_ for each period. The mean and 95% CrI (in parentheses) are labeled below or above the violin plots. (C) Prediction using parameters from period 4 (February 2-16). (D) Prediction using parameters from period 3 (January 23-February 1). (E) Prediction using parameters from period 2 (January 10-22). The shaded areas in (A, C, D and E) are 95% CrI. (F) Estimated number of active infectious cases in Wuhan from January 1 to March 8.

Strikingly, we estimated low ascertainment rates across periods, which were 0.15 (0.13-0.17) for the first two periods, and 0.14 (0.12-0.17), 0.10 (0.08-0.12), and 0.16 (0.13-0.21) for the other three periods, respectively (**Extended Data Table 4**). Even with the universal community symptom screening implemented on February 17 to 19, the ascertainment rate was only increased to 0.16. Based on the fitted model using data from January 1 to February 29, we projected the cumulative number of ascertained cases to be 32,562 (30,234-34,999) by March 8, close to the actual reported number of 32,583. This was equivalent to an overall ascertainment rate of 0.13 (0.11-0.16) given the estimated total infections of 248,022 (199,474-302,464). The model also projected that the number of daily active infections in Wuhan, including both ascertained and unascertained, peaked at 55,651 (45,204-67,834) on February 2 and dropped afterwards to 683 (437-1,003) on March 8 (**Fig. 2F**). If the trend remained unchanged, the number of ascertained infections would first become zero on March 25 (95% CrI: March 18 to April 2), while the clearance of all infections would occur on April 21 (April 8 to May 11), 2020 (**Extended Data Table 7**). The first day of zero ascertained case in Wuhan was reported on March 18, which was on the lower 95% CrI of our prediction, indicating that control measures might have been enhanced in March.

The large fraction of unascertained cases has important implications for continuing surveillance and interventions.^18^ Based on stochastic simulations, we estimated the probability of resurgence after lifting all controls, assuming the transmission rate, ascertainment rate, and daily population movement were resumed to values of the first period (**Methods**). Because of the latent, presymptomatic, and unascertained cases, the source of infection would not be completely cleared shortly after the first day of zero ascertained cases. We found that if control measures were lifted 14 days after the first day of zero ascertained cases, despite sparse new cases might be ascertained during the observation period, the probability of resurgence could be as high as 0.97, and the surge was predicted to occur on day 36 (95% CrI: 28-48) after lifting controls (**Fig. 3**). If we were to impose a more stringent criterion of lifting controls after observing no ascertained cases in a consecutive period of 14 days, the probability of resurgence would drop to 0.33, with possible resurgence delayed to day 43 (95% CrI: 34-58) after lifting controls (**Fig. 3**). These results highlighted the risk of ignoring unascertained cases in switching intervention strategies, despite using an over-simplified model without considering other factors such as imported cases, changes in temperature and humidity, and a stepwise lifting strategy that is currently adopted by Wuhan and other cities in China.

**Fig. 3.**
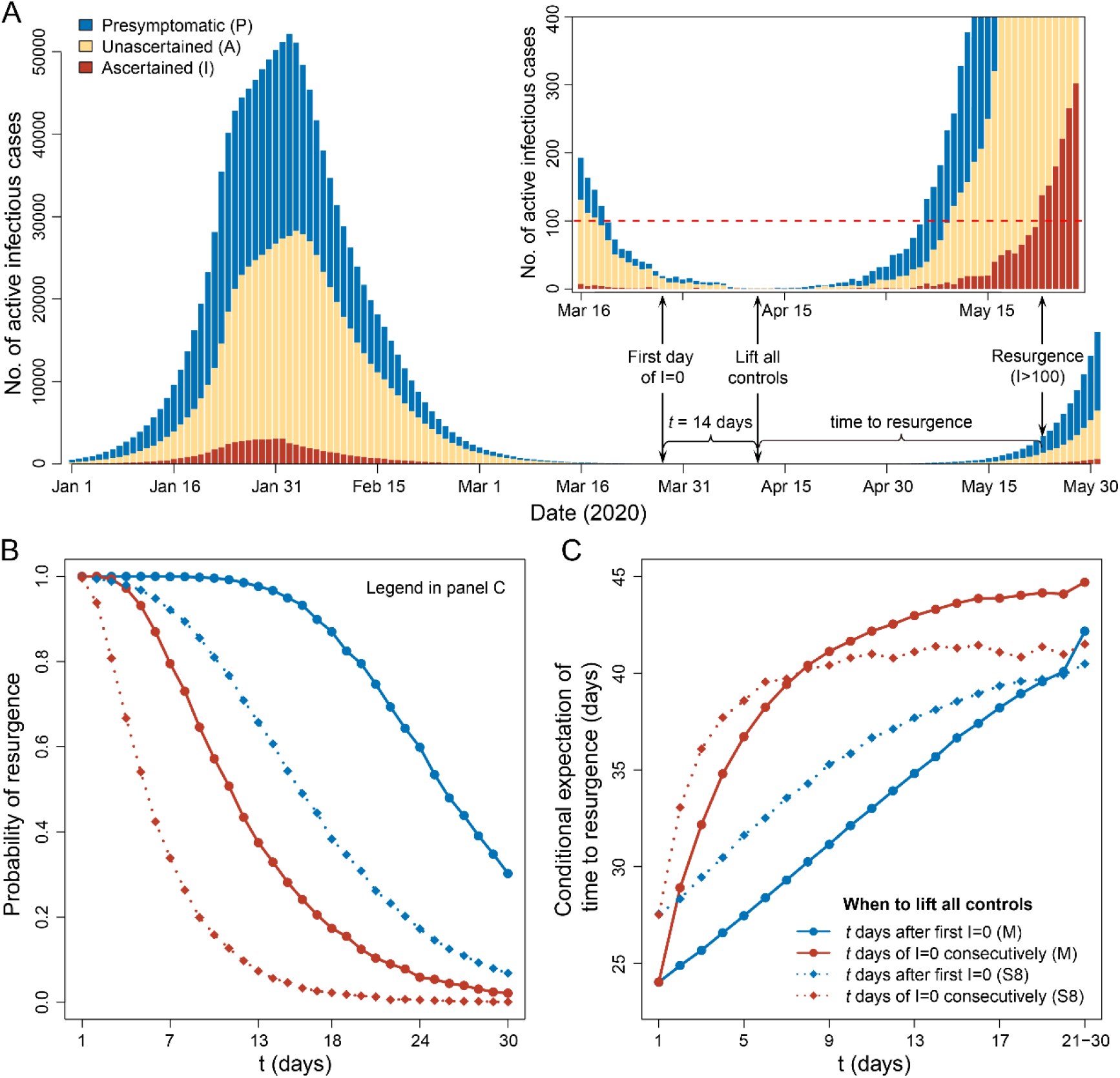
Risk of resurgence after lifting controls. We considered the main model (M) and the sensitivity analysis model S8 (see **Methods**). In model M, we assumed the initial ascertainment rate *r*_0_ = 0.23, and thus had a 0.13 estimate of the overall ascertainment rate. In model S8, we assumed no unascertained cases in the initial state and thus had a 0.46 estimate of the overall ascertainment rate. For each model, we simulated epidemic curves based on 10,000 sets of parameter values from MCMC, assuming transmission rate *b*, ascertainment rate *r*, and population movement *n* were resumed to values before *Chunyun* after lifting controls. A resurgence was defined by when the number of active ascertained infections raised to over 100. (A) Illustration of a simulated curve under the main model with control measures lifted 14 days after the first day of no ascertained cases. The inserted panel is a zoom-in plot from March 16 to May 28. (B) Probability of resurgence if control measures were lifted *t* days after the first observation of no ascertained cases, or after observing zero ascertained cases in a consecutive period of *t* days. (C) Expectation of time to resurgence conditional on the occurrence of resurgence. We grouped the last 10 days (*t* = 21 to 30) to calculate the expected time to resurgence because of their low probability of resurgence.

We performed a series of sensitivity analyses to test the robustness of our results by smoothing the outlier data point on February 1, varying lengths of latent and infectious periods, duration from symptom onset to isolation, ratio of transmissibility of presymptomatic/unascertained cases to ascertained cases, and initial ascertainment rate (**Extended Data Tables 4-7, Extended Data Figs. 4-11**). Our major findings of remarkable decrease in *R*_0_ after interventions and the existence of substantial unascertained cases was robust in all sensitivity analyses. Consistent with simulation results, the estimated ascertainment rates were positively correlated with the specified initial ascertainment rate. When we specified the initial ascertainment rate as 0.14 or 0.42, the estimated overall ascertainment rate would be 0.08 (0.07-0.10) and 0.23 (0.19-0.28), respectively (**Extended Data Table 4, Extended Data Figs. 9-10**). If we assumed an extreme scenario with no unascertained cases in the early outbreak (model S8; **Extended Data Fig. 11**), the estimated ascertainment rate would be 0.47 (0.38-0.57) overall and 0.58 (0.45-0.73) for the last period, which would represent an upper bound of the ascertainment rate. In this model, because of the higher ascertainment rate compared to the main analysis, we estimated a lower probability of resurgence of 0.06 when lifting controls after 14 days of no ascertained cases, and a longer time to resurgence, occurring on day 41 (95% CrI: 31-56) after lifting controls (**Fig. 3**). We also tested a simplified model assuming complete ascertainment anytime, but this simplified model performed significantly worse than the full model, especially in fitting the rapid growth before interventions (**Extended Data Fig. 12**).

Our finding of a large fraction of unascertained cases, despite the strong surveillance in Wuhan, indicated the existence of many asymptomatic or mild-symptomatic but infectious cases during the outbreak, highlighting a key challenge to the COVID-19 epidemic control.^19^ There is accumulating evidence on the existence of many asymptomatic cases. For example, asymptomatic cases were estimated to account for 18% of the infections onboard the Diamond Princess Cruise ship^7^ and 31% of the infected Japanese evacuated from Wuhan.^8^ In addition, it was reported that 29 of the 33 (88%) infected pregnant women were asymptomatic by universal screening of 210 women admitted for delivery between March 22 and April 4 in New York City.^9^ Several reports also highlighted the difficulty in detecting COVID-19 cases: about two thirds of the cases exported from mainland China remained undetected worldwide,^20^ and the detection capacity varied from 11% in low surveillance countries to 40% in high surveillance countries.^21,22^ By modeling the epidemics in other cities, it was also estimated that the ascertainment rate of infected individuals was about 24.4% in China (excluding Hubei province)^14^ and 14% in Wuhan prior to travel ban.^15^ Consistent with these studies, our extensive analyses of the most comprehensive epidemic data from Wuhan also indicated an overall ascertainment rate between 7% and 28% (**Extended Data Table 4**, excluding the extreme scenario of model S8). These results were also consistent with emerging serological studies, showing much higher seroprevalence than the reported case prevalence in different regions of the world.^23,24^ The large fraction of unascertained cases would lead to about one month delay between the first occurrence of no ascertained cases and the clearance of all infections (**Extended Data Table 6**), imposing a high risk of resurgence after lifting controls (**Fig. 3**). Therefore, understanding the proportion of unascertained cases and the asymptomatic transmissibility will be critical for prioritization of the surveillance and control measures.^18,25^ Currently, Wuhan is implementing a strategy to normalize and restore societal activities gradually while maintaining strong disease surveillance. The experience and outcome of Wuhan will be valuable to other countries who will eventually face the same issue.

In the absence of interventions, *R*_0_ is the basic reproduction number, which is a key measurement of the virus transmissibility. We noted that our *R*_0_ estimate of 3.54 (3.41-3.66) before any interventions was at the higher end of the range of *R*_0_ estimated by other studies using early epidemic data from Wuhan (1.40-6.49 with a median of 2.79).^2,14,15,26-28^ Several plausible reasons might explain the discrepancy, including potential impact of unascertained cases, more complete case records in our analysis, and different time periods analyzed. If we considered a model starting from the first COVID-19 case reported in Wuhan (**Extended Data Fig. 13**), from which we estimated a lower *R*_0_ of 3.38 (3.28-3.48) before January 23, 2020, similar to the value of 3.15 reported by a recent study.^27^ Nevertheless, this reproduction number was still much higher than the earlier estimates and those for SARS and MERS,^29,30^ featuring another challenge to control the spread of COVID-19.

Taken together, our modeling study delineated the full-spectrum dynamics of the COVID-19 outbreak in Wuhan, and highlighted two key features of the outbreak: a high proportion of asymptomatic or mild-symptomatic cases who were difficult to detect, and high transmissibility. These two features synergistically propelled the global pandemic of COVID-19, imposing grand challenges to control the outbreak. Nevertheless, lessons from Wuhan have demonstrated that vigorous and multifaceted containment efforts can considerably control the size of the outbreak, as evidenced by the remarkable decrease of *R*_0_ from 3.54 to 0.27 and an estimated 96.1% reduction of infections till March 8. These are important information for other countries combatting the outbreak.

Some limitations of our study should be noted. First, we need field investigations and serologic studies to confirm our estimate of the ascertainment rate, and the generalizability to other places is unknown. This may depend on the detection capacity in different locations.^22^ Second, due to the delay in laboratory tests, we might have missed some cases and therefore underestimated the ascertainment rate, especially for the last period. Third, we excluded clinically diagnosed cases without laboratory confirmation to reduce false positive diagnoses, which, however, would lead to lower estimates of ascertainment rates, especially for the third and fourth periods when many clinically diagnosed cases were reported.^4^ The variation in the estimated ascertainment rates across periods reflected a combined effect of the evolving surveillance, interventions, medical resources, and case definitions across time periods.^4,31^ Fourth, our model assumed homogeneous transmission within the population while ignoring heterogeneity between groups by sex, age, geographic regions and socioeconomic status. Furthermore, individual variation in infectiousness, such as superspreading events,^32^ is known to result in a higher probability of stochastic extinction given a fixed population *R*_0_.^33^. Therefore, we might have overestimated the probability of resurgence.

Finally, we could not evaluate the impact of individual interventions based on the epidemic curve from a single city, because many interventions were applied simultaneously. Future work by modeling heterogeneous transmission between different groups and joint analysis with data from other cities will lead to deeper insights on the effectiveness of different control strategies.^27,34^

## Data Availability

Data will be released once granted by the authorities.

## Data and codes availability

R codes and data are available at http://chaolongwang.github.io/codes_covid19.zip.

## Acknowledgements

We thank Dr. Huaiyu Tian from Beijing Normal University and the anonymous reviewers for constructive comments. This study was partly supported by the Fundamental Research Funds for the Central Universities (2019kfyXMBZ015), the 111 Project (X.H., S.C., D.W., C.W., T.W.). X.L. is supported by Harvard University.

## Author contributions

TW, XL and CW designed the study. XH, SC, XL and CW developed statistical methods. XH, SC, and DW performed data analysis. CW wrote the first draft of the manuscript. All authors reviewed and edited the manuscript.

## Methods

### Data of COVID-19 cases in Wuhan

Detailed description of the data can be found in Pan *et al*.^4^ Briefly, information of COVID-19 cases from December 8, 2019 till March 8, 2020 were extracted from the municipal Notifiable Disease Report System on March 9, 2020. Date of symptom onset (the self-reported date of symptoms such as fever, cough, or other respiratory symptoms) and date of confirmed diagnosis were collected for each case. For the consistency of case definition throughout the periods, we only included 32,583 laboratory-confirmed cases who were tested positive for SARS-CoV-2 by the real-time reverse-transcription-polymerase-chain-reaction (RT-PCR) assay or high-throughput sequencing of nasal and pharyngeal swab specimens.

### Estimation of ascertainment rate using cases exported to Singapore

As of May 10, 2020, a total of 24 confirmed COVID-19 cases in Singapore were reported to import from China, among which 16 were imported from Wuhan before the *cordons sanitaire* on January 23 (**Extended Data Table 1**). Based on VariFlight Data (https://data.variflight.com/en/), the total number of passengers from Wuhan to Singapore between January 18 and 23, 2020 was 2,722. Therefore, the cumulative infection rate among the passengers was 0.59% (=16/2722, 95% CI: 0.30-0.88%). These cases had symptom onset from January 21 to 30, 2020. In Wuhan, a total of 12,433 confirmed cases were reported to have symptom onset in the same period, equivalent to a cumulative infection rate of 0.124% (95% CI: 0.122–0.126%) by assuming a population size of 10 million for Wuhan. By further assuming complete ascertainment of early cases in Singapore, which is well known for excellent surveillance strength,^21,22^ the ascertainment rate in Wuhan was estimated to be 0.23 (95% CI: 0.14–0.42), corresponding to 0.77 (95% CI: 0.58-0.86) of the infections being unascertained during the early outbreak in Wuhan. This represents a conservative estimate for two reasons: (1) the assumption of perfect ascertainment in Singapore ignored potential asymptomatic cases;^7,8^ and (2) the number of imported cases with onset between January 21 and 30 was censored due to suspension of flights after Wuhan lockdown. We used these results to set the initial value and the prior distribution of ascertainment rates in our model.

### The SAPHIRE model

We extended the classic susceptible-exposed-infectious-recovered (SEIR) model to a SAPHIRE model (**Fig. 1**), which incorporates three additional compartments to account for presymptomatic infectiousness (*P*), unascertained cases (*A*), and case isolation in the hospital (*H*). We chose to analyze data from January 1, 2020, when the Huanan Seafood Market was disinfected, and thus did not model the zoonotic force of infection. We assumed a constant population size *N* = 10,000,000 with equal number of daily inbound and outbound travelers *n*, where *n =* 500,000 for January 1-9, 800,000 for January 10-22 due to *Chunyun*, and 0 after *cordons sanitaire* from January 23.^3^ We divided the population into *S* susceptible, *E* exposed, *P* presymptomatic infectious, *A* unascertained infectious, *I* ascertained infectious, *H* isolated, and *R* removed individuals. Dynamics of these seven compartments across time *t* were described by the following set of ordinary differential equations:

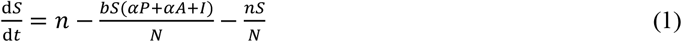

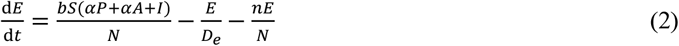

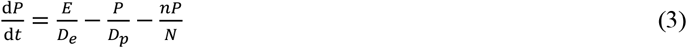

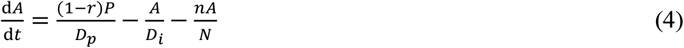

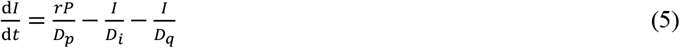

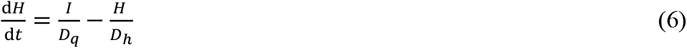

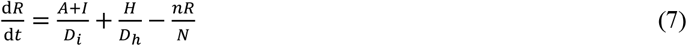

where *b* was the transmission rate, defined as the number of individuals that an ascertained case can infect per day; *α* was the ratio of the transmission rate of presymptomatic/unascertained over ascertained cases; *r* was ascertainment rate; *D_e_* was the latent period; *D_p_* was the presymptomatic infectious period; *D_i_* was the symptomatic infectious period; *D_q_* was the duration from illness onset to isolation; and *D_h_* was the isolation period in hospital. The effective reproduction number *R*_0_ could be computed as

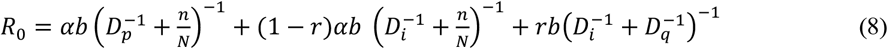

where the three terms represent infections contributed by presymptomatic, unascertained, and ascertained cases, respectively. We adjusted the infectious periods of each type of cases by taking population movement 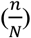 and isolation 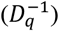 into account.

### Parameter settings and initial states

Parameter settings for the main analysis were summarized in **Extended Data Table 2**. We set *α* = 0.55 according to Li *et al*.^15^, assuming lower transmissibility for presymptomatic and unascertained cases. We assumed an incubation period of 5.2 days and a presymptomatic infectious period of *D_p_* = 2.3 days.^2,10^ Thus the latent period was *D_e_* = 5.2 − 2.3 = 2.9 days. Because presymptomatic infectiousness was estimated to account for 44% of the total infections of ascertained cases,^10^ we set the total infectious period as 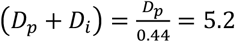, thus the symptomatic infectious period was *D_i_* = 2.9 days. We 8 set a long isolation period of *D_h_* = 30 days, but this parameter has no impact on our fitting procedure and the final parameter estimates. The duration from symptom onset to isolation was estimated to be *D_q_* = 21, 15, 10, 6, and 2 days as the median time length from onset to confirmed diagnosis in each period, respectively.

Based on the settings above, we specified the initial state of the model on December 31, 2019 (**Extended Data Table 3**). The initial number of ascertained symptomatic cases *I* (0) was specified as the number of ascertained cases with onset during December 29-31, 2019. We assumed the initial ascertainment rate was *r*_0_, and thus the initial number of unascertained cases was 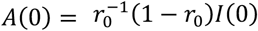. We denoted *P_I_* (0) and *E_I_* (0) as the numbers of ascertained cases with onset during January 1-2, 2020 and during January 3-5, 2020, respectively. Then, the initial numbers of exposed cases 18 and presymptomatic cases were set as 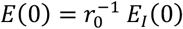 and 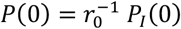, respectively. We assumed *r*_0_ = 0.23 in our main analysis based on the point estimate using the Singapore data (described 20 above).

### Estimation of parameters in the SEIR model

Considering the time-varying strength of control measures, we assumed *b* = *b*_12_ and *r* = *r*_12_ for the first two periods, *b* = *b*_3_ and *r* = *r*_3_ for period 3, *b* = 4 and *r* = *r*_4_ for period 4, and *b* = *b*_5_ and *r* = *r*_5_ for period 5. We assumed the observed number of ascertained cases with symptom onset on day *d*, denoted as *x_d_*, followed a Poisson distribution with rate 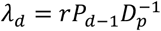, where *P_d_*_−1_ was the 27 expected number of presymptomatic cases on day (*d*−1). We fit the observed data from January 1 to 28 February 29 (*d* = 1,2, …, *D*, and *D* = 60) and used the fitted model to predict the trend from March 1 to 8. Thus, the likelihood function was

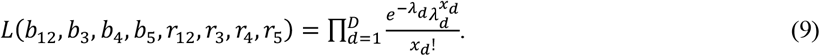

We estimated *b*_12_, *b*_3_, *b*_4_, *b*_5_ *r*_12_, *r*_3_, *r*_4_, and *r*_5_ by Markov Chain Monte Carlo (MCMC) with the Delayed Rejection Adaptive Metropolis (DRAM) algorithm implemented in the R package *BayesianTools* (version 0.1.7).^35^ We used a non-informative flat prior of Uinf(0,2) for *b*_12_, *b*_3_, *b*_4_, and *b*_5_. For *r*_12_, we used an informative prior of Beta(7.3,24.6) by matching the first two moments of the estimate using Singapore data (described above). We reparameterized *r*_3_, *r*_4_, and *r*_5_ by

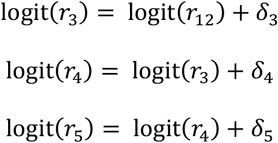

where logit 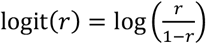. In the MCMC, we sampled *δ*_3_, *δ*_4_, and *δ*_5_ from the prior of *N* (0,1). We set a burn-in period of 40,000 iterations and continued to run 100,000 iterations with a sampling step size of 10 iterations. We repeated MCMC with three different sets of initial values and assessed the convergence by the trace plot and the multivariate Gelman-Rubin diagnostic (**Extended Data Fig. 3**).^36^ Estimates of parameters were presented as posterior means and 95% credible intervals (CrIs) from 10,000 MCMC samples. All the analyses were performed in R (version 3.6.2) and the Gelman-Rubin diagnostic was calculated using the *gelman.diag* function in the R package *coda*.

### Stochastic simulations

We used stochastic simulations to obtain 95% CrI of fitted/predicted epidemic curve. Given a set of parameter values from MCMC, we performed the following multinomial random sampling:

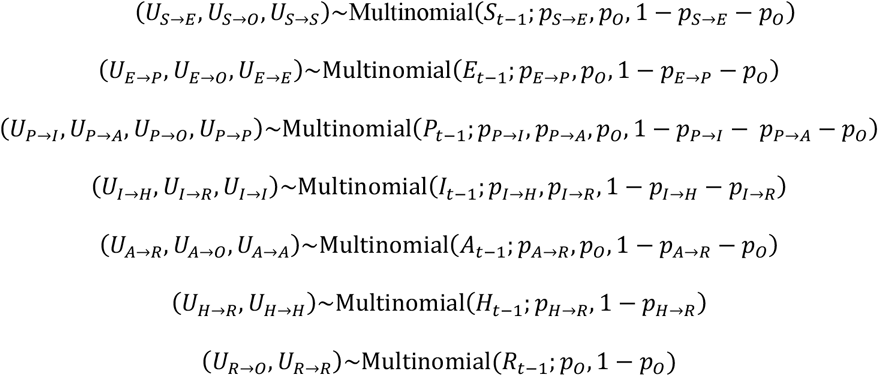

where *O* denotes the status of outflow population, *p_o_* = *nN*^−1^ denotes the outflow probability, and other quantities are status transition probabilities, including *p_S_*_→_*_E_* = *b*(*αp_t_*_−1_ + *αA_t_*_−1_ + *I_t_*_−1_)*N*^−1^, 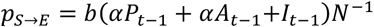, 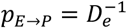, 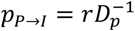, 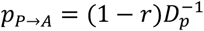, 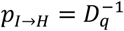, and 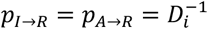. The SAPHIRE model described by Eqs. 1-7 is equivalent to the following stochastic dynamics:

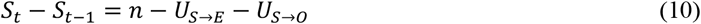

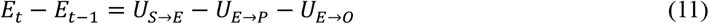

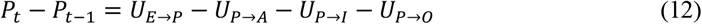

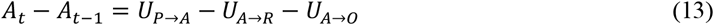

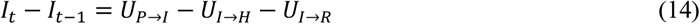

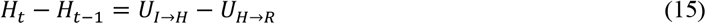

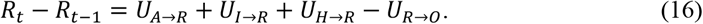

We repeated the stochastic simulations for all 10,000 sets of parameter values sampled by MCMC to construct the 95% CrI of the epidemic curve by the 2.5 and 97.5 percentiles at each time point.

### Prediction of epidemic ending date and the risk of resurgence

Using the stochastic simulations described above, we predicted the first day of no new ascertained cases and the date of clearance of all active infections in Wuhan, assuming continuation of the same control measures as the last period (*i.e*., same parameter values).

We also evaluated the risk of outbreak resurgence after lifting control measures. We considered lifting all controls (1) at *t* days after the first day of zero ascertained cases, or (2) after a consecutive period of *t* days with no ascertained cases. After lifting controls, we set the transmission rate *b*, ascertainment rate *r*, and population movement *n* to be the same as the first period, and continued the stochastic simulation to stationary state. Time to resurgence was defined as the number of days from lifting controls to when the number of ascertained cases *I* reached 100. We performed 10,000 simulations with 10,000 sets of parameter values sampled from MCMC (as described above). We calculated the probability of resurgence as the proportion of simulations in which a resurgence occurred, as well as the time to resurgence conditional on the occurrence of resurgence.

### Simulation study for method validation

To validate our method, we performed two-period stochastic simulations (Eqs. 10-16) with transmission rate *b* = *b*_1_=1.27, ascertainment rate *r* = *r*_1_ = 0.2, daily population movement *n* = 500,000, and duration from illness onset to isolation *D_q_* = 20 days for the first period (so that *R*_0_ = 3.5 according to Eq. 8), and *b* = *b*_2_ = 0.41, *r* = *r*_2_ = 0.4, *n* = 0, and *D_q_* = 5 for the second period (so that *R*_0_ = 1.2 according to Eq. 8). Lengths of both periods were set to 15 days, and the initial ascertainment rate was set to *r*_0_ = 0.3, while the other parameters and initial states were set as those in our main analysis (**Extended Data Tables 2-3**). We repeated stochastic simulations 100 times to generate 100 datasets. For each dataset, we applied our MCMC method to estimate *b*_1_, *b*_2_, *r*_1_ and *r*_2_, while setting all other parameters and initial values the same as the true values. We translated *b*_1_ and *b*_2_ into (*R*_0_)_1_ and (*R*_0_)_2_ according to Eq. 8, and focused on evaluating the estimates of (*R*_0_)_1_, (*R*_0_)_2_, *r*_1_ and *r*_2_. We also tested the robustness to misspecification of the latent period *D_e_*, presymptomatic infectious period *D_p_*, symptomatic infectious period *D_i_*, duration from illness onset to isolation *D_q_*, ratio of transmissibility between unascertained/presymptomatic cases and ascertained cases *α*, and initial ascertainment rate *r*_0_. In each test, we changed the specified value of a parameter (or initial state) to be 20% lower or higher than its true value, while keeping all other parameters unchanged. When we changed the value of *r*_0_, we adjusted the initial states *A*(0), *P*(0), and *E*(0) according to **Extended Data Table 3**.

For each simulated dataset, we ran the MCMC method with 20,000 burn-in iterations and an additional 30,000 iterations. We sampled parameter values from every 10 iterations, resulting in 3,000 MCMC samples. We took the mean across 3,000 MCMC samples as the final estimates and displayed results for 100 repeated simulations using boxplots.

### Sensitivity analyses for the real data

We designed nine sensitivity analyses to test the robustness of our real data results. For each of the sensitivity analyses, we fixed parameters and initial states to be the same as the main analysis except for those mentioned below.

(S1) Adjust the reported incidences from January 29 to February 1 to their average. We suspect the spike of incidences on February 1 might be caused by approximate-date records among some patients admitted to the centralized quarantine after February 2. The actual illness onset dates for these patients were likely to be between January 29 and February 1.
(S2) Assume an incubation period of 4.1 days (lower 95% CI from reference^2^) and presymptomatic infectious period of 1.1 days (lower 95% CI from reference^10^ is 0.8 days, but our discrete stochastic model requires *D_p_* > 1), equivalent to set *D_e_* = 3 and *D_p_* = 1.1, and adjust *P*(0) and *E*(0) accordingly.
(S3) Assume an incubation period of 7 days (upper 95% CI from reference 2) and presymptomatic infectious period of 3 days (upper 95% CI from reference 10), equivalent to set *D_e_* = 4 and *D_p_* = 3, and adjust *P*(0) and *E*(0) accordingly.
(S4) Assume the transmissibility of the presymptomatic and unascertained cases is *α* = 0.46 (lower 95% CI from reference^15^) of the ascertained cases.
(S5) Assume the transmissibility of the presymptomatic and unascertained cases is *α* = 0.62 (upper 95% CI from reference^15^) of the ascertained cases.
(S6) Assume the initial ascertainment rate is *r*_0_ = 0.14 (lower 95% CI of the estimate using Singapore data) and adjust *A*(0), *P*(0), and *E*(0) accordingly.
(S7) Assume the initial ascertainment rate is *r*_0_ = 0.42 (upper 95% CI of the estimate using Singapore data) and adjust *A*(0), *P*(0), and *E*(0) accordingly.
(S8) Assume the initial ascertainment rate is *r*_0_ = 1 (theoretical upper limit) and adjust *A*(0), *P*(0), and *E*(0) accordingly.
(S9) Assume no unascertained cases by fixing *r*_0_ = *r*_12_ = *r*_3_ = *r*_4_ = *r*_5_ = 1. We test if the full model is significantly better than this simplified model using likelihood ratio test.

**Extended Data Table 1.** COVID-19 cases exported from Wuhan to Singapore before January 23, 2020.

| Case ID | Arrival date | Symptom onset | Confirmed date |
| --- | --- | --- | --- |
| 1 | 2020/1/20 | 2020/1/21 | 2020/1/23 |
| 2 | 2020/1/21 | 2020/1/21 | 2020/1/24 |
| 3 | 2020/1/20 | 2020/1/23 | 2020/1/24 |
| 4 | 2020/1/22 | 2020/1/23 | 2020/1/25 |
| 5 | 2020/1/18 | 2020/1/24 | 2020/1/27 |
| 6 | 2020/1/19 | 2020/1/25 | 2020/1/27 |
| 7 | 2020/1/23 | 2020/1/24 | 2020/1/27 |
| 8 | 2020/1/19 | 2020/1/24 | 2020/1/28 |
| 9 | 2020/1/19 | 2020/1/24 | 2020/1/29 |
| 10 | 2020/1/20 | 2020/1/21 | 2020/1/29 |
| 11 | 2020/1/22 | 2020/1/27 | 2020/1/29 |
| 12 | 2020/1/22 | 2020/1/26 | 2020/1/29 |
| 13 | 2020/1/21 | 2020/1/28 | 2020/1/30 |
| 16 | 2020/1/22 | 2020/1/23 | 2020/1/31 |
| 18 | 2020/1/22 | 2020/1/30 | 2020/2/1 |
| 26 | 2020/1/21 | 2020/1/28 | 2020/2/4 |

Source: https://co.vid19.sg/singapore/dashboard

**Extended Data Table 2.** Parameter settings for five periods in the main analysis.

| Parameter | Meaning | Jan 1-9 | Jan 10-22 | Jan 23-Feb 1 | Feb 2-16 | Feb 17-Mar 8 |
| --- | --- | --- | --- | --- | --- | --- |
| $b$ | Transmission rate of ascertained cases | $b_{12}$ | $b_{12}$ | $b_3$ | $b_4$ | $b_5$ |
| $r$ | Ascertainment rate | $r_{12}$ | $r_{12}$ | $r_3$ | $r_4$ | $r_5$ |
| $\alpha$ | Ratio of transmission rate for presymptomatic and unascertained over ascertained cases | 0.55 | 0.55 | 0.55 | 0.55 | 0.55 |
| $D_e$ | Latent period | 2.9 | 2.9 | 2.9 | 2.9 | 2.9 |
| $D_p$ | Presymptomatic infectious period | 2.3 | 2.3 | 2.3 | 2.3 | 2.3 |
| $D_i$ | Symptomatic infectious period | 2.9 | 2.9 | 2.9 | 2.9 | 2.9 |
| $D_q$ | Duration from illness onset to isolation | 21 | 15 | 10 | 6 | 2 |
| $D_h$ | Isolation period | 30 | 30 | 30 | 30 | 30 |
| $N$ | Population size | 10,000,000 | 10,000,000 | 10,000,000 | 10,000,000 | 10,000,000 |
| $n$ | Daily inbound and outbound size | 500,000 | 800,000 | 0 | 0 | 0 |

**Extended Data Table 3.** Initial state of the model for the main analysis.

| Variable | Meaning | Value | Note |
| --- | --- | --- | --- |
| $S(0)$ | Number of susceptible individuals | 9,999,021 | $S = N - E - P - A - I - H - R$ |
| $E(0)$ | Number of exposed cases | 478 | $E(0) = r_0^{-1}E_I(0)$, where $E_I(0)$ was the number of ascertained cases with onset during Jan 3-5, 2020 (day $(D_p + 1)$ to day $(D_p + D_e)$) * |
| $P(0)$ | Number of presymptomatic cases | 326 | $P(0) = r_0^{-1}P_I(0)$, where $P_I(0)$ was the number of ascertained cases with onset during Jan 1-2, 2020 (day 1 to day $D_p$) * |
| $I(0)$ | Number of ascertained cases | 34 | Number of ascertained cases with onset during Dec 29-31, 2019 ($D_i$ days before day 1) |
| $A(0)$ | Number of unascertained cases | 114 | $A(0) = r_0^{-1}(1 - r_0)I(0)$ * |
| $H(0)$ | Number of isolated cases | 27 | Number of cases reported by Dec 31, 2019 |
| $R(0)$ | Number of removed individuals | 0 | Number of cases recovered by Dec 31, 2019 |

\* The initial ascertainment rate $r_0$ was assumed to be 0.23 in the main analysis. Day 1 was January 1, 2020.

**Extended Data Table 4.** Estimated ascertainment rates from the main and sensitivity analyses.

| **Analysis** | **$r_{12}$** | **$r_3$** | **$r_4$** | **$r_5$** | **Overall** |
| --- | --- | --- | --- | --- | --- |
| Main | 0.15 (0.13-0.17) | 0.14 (0.12-0.17) | 0.10 (0.08-0.12) | 0.16 (0.13-0.21) | 0.13 (0.11-0.16) |
| S1 | 0.15 (0.12-0.17) | 0.15 (0.12-0.18) | 0.11 (0.09-0.14) | 0.18 (0.14-0.24) | 0.14 (0.11-0.17) |
| S2 | 0.14 (0.12-0.17) | 0.15 (0.12-0.18) | 0.11 (0.08-0.13) | 0.17 (0.13-0.22) | 0.14 (0.11-0.17) |
| S3 | 0.14 (0.12-0.16) | 0.13 (0.10-0.16) | 0.09 (0.07-0.11) | 0.16 (0.12-0.20) | 0.12 (0.10-0.15) |
| S4 | 0.15 (0.12-0.17) | 0.14 (0.11-0.17) | 0.10 (0.08-0.12) | 0.16 (0.12-0.21) | 0.13 (0.11-0.16) |
| S5 | 0.15 (0.13-0.17) | 0.14 (0.11-0.17) | 0.10 (0.08-0.12) | 0.16 (0.13-0.21) | 0.13 (0.11-0.16) |
| S6 | 0.09 (0.08-0.10) | 0.09 (0.07-0.10) | 0.06 (0.05-0.08) | 0.10 (0.08-0.13) | 0.08 (0.07-0.10) |
| S7 | 0.26 (0.22-0.30) | 0.25 (0.20-0.30) | 0.17 (0.14-0.22) | 0.29 (0.22-0.38) | 0.23 (0.19-0.28) |
| S8 | 0.54 (0.47-0.62) | 0.50 (0.41-0.59) | 0.35 (0.28-0.43) | 0.58 (0.45-0.73) | 0.47 (0.38-0.57) |

The estimates were displayed as mean (95% CrI) based on 10,000 MCMC samples.

**Extended Data Table 5.** Estimated transmission rates from the main and sensitivity analyses.

| **Analysis** | **$b_{12}$** | **$b_3$** | **$b_4$** | **$b_5$** |
| --- | --- | --- | --- | --- |
| Main | 1.31 (1.25-1.37) | 0.40 (0.38-0.42) | 0.17 (0.16-0.19) | 0.10 (0.08-0.12) |
| S1 | 1.31 (1.25-1.38) | 0.37 (0.34-0.39) | 0.17 (0.16-0.18) | 0.10 (0.08-0.12) |
| S2 | 1.50 (1.43-1.58) | 0.53 (0.51-0.56) | 0.25 (0.24-0.27) | 0.15 (0.13-0.18) |
| S3 | 1.46 (1.39-1.54) | 0.34 (0.31-0.36) | 0.11 (0.10-0.13) | 0.04 (0.03-0.06) |
| S4 | 1.53 (1.46-1.61) | 0.47 (0.44-0.50) | 0.21 (0.19-0.22) | 0.12 (0.1-0.14) |
| S5 | 1.18 (1.13-1.23) | 0.36 (0.34-0.38) | 0.16 (0.15-0.17) | 0.09 (0.07-0.10) |
| S6 | 1.34 (1.28-1.40) | 0.41 (0.38-0.44) | 0.18 (0.17-0.19) | 0.10 (0.08-0.12) |
| S7 | 1.27 (1.21-1.34) | 0.39 (0.36-0.41) | 0.17 (0.16-0.18) | 0.10 (0.08-0.12) |
| S8 | 1.20 (1.14-1.27) | 0.36 (0.34-0.39) | 0.17 (0.16-0.18) | 0.10 (0.09-0.12) |
| S9 | 0.93 (0.92-0.94) | 0.26 (0.25-0.27) | 0.17 (0.16-0.17) | 0.18 (0.16-0.20) |

The estimates were displayed as mean (95% CrI) based on 10,000 MCMC samples.

**Extended Data Table 6.**
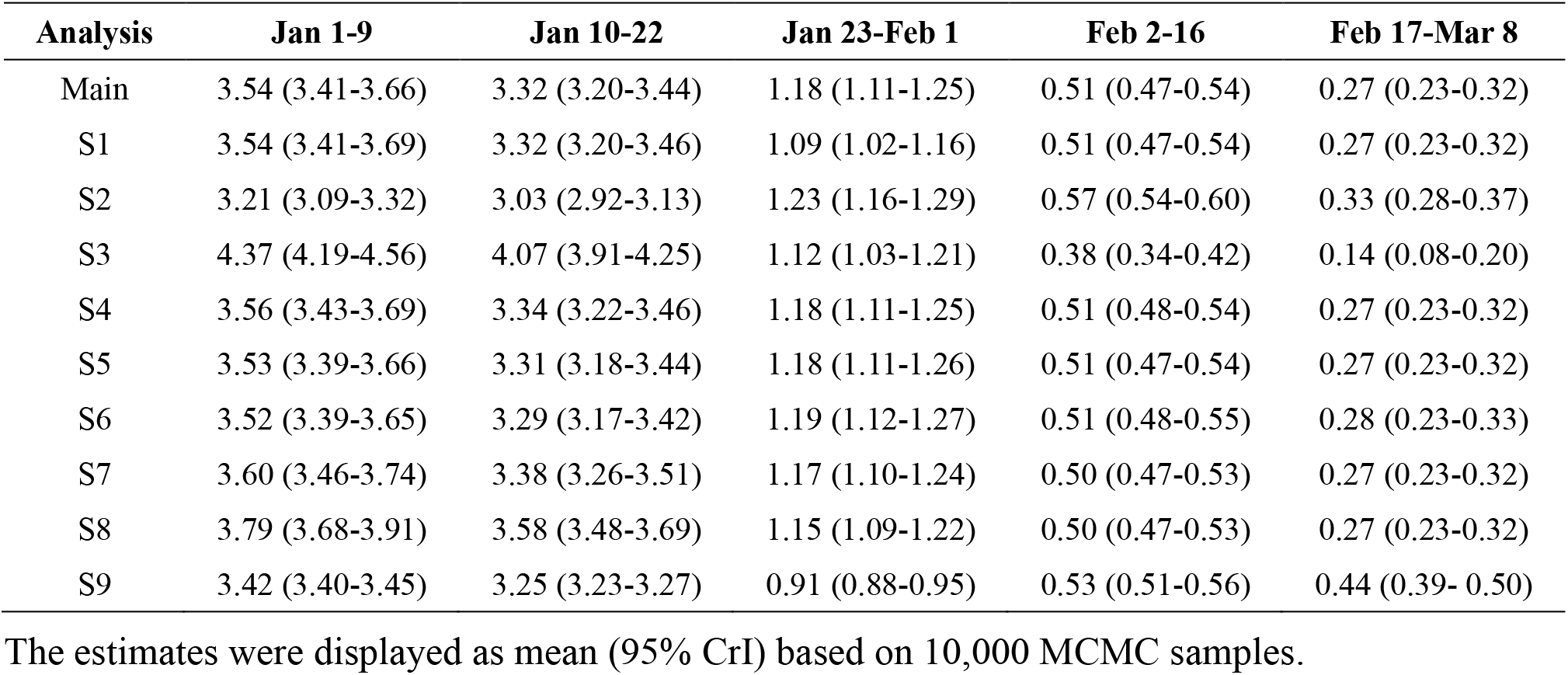
Estimated *R*_0_ for different periods from the main and sensitivity analyses.

**Extended Data Table 7.** Prediction of the ending date of COVID-19 epidemic in Wuhan from the main and sensitivity analyses.

| Analysis | First day of no ascertained infections | Clearance of all infections |
| --- | --- | --- |
| Main | Mar 25 (Mar 18 to Apr 2) | Apr 21 (Apr 8 to May 11) |
| S1 | Mar 25 (Mar 18 to Apr 1) | Apr 20 (Apr 7 to May 11) |
| S2 | Mar 26 (Mar 19 to Apr 2) | Apr 22 (Apr 8 to May 13) |
| S3 | Mar 23 (Mar 16 to Mar 31) | Apr 19 (Apr 6 to May 8) |
| S4 | Mar 25 (Mar 18 to Apr 1) | Apr 21 (Apr 8 to May 11) |
| S5 | Mar 25 (Mar 18 to Apr 2) | Apr 21 (Apr 8 to May 11) |
| S6 | Mar 25 (Mar 18 to Apr 2) | Apr 24 (Apr 11 to May 15) |
| S7 | Mar 25 (Mar 18 to Apr 1) | Apr 17 (Apr 3 to May 7) |
| S8 | Mar 24 (Mar 17 to Mar 31) | Apr 10 (Mar 28 to Apr 29) |
| S9 | Apr 2 (Mar 24 to Apr 13) | Apr 19 (Apr 2 to May 15) |

The estimates were displayed as mean date (95% CrI) based on 10,000 stochastic simulations with parameter values from MCMC sampling.
First day of no ascertained infections means the first day of $I = 0$ .
Clearance of all infections means the first day of $E = P = A = I = 0$ .

**Extended Data Fig. 1.**
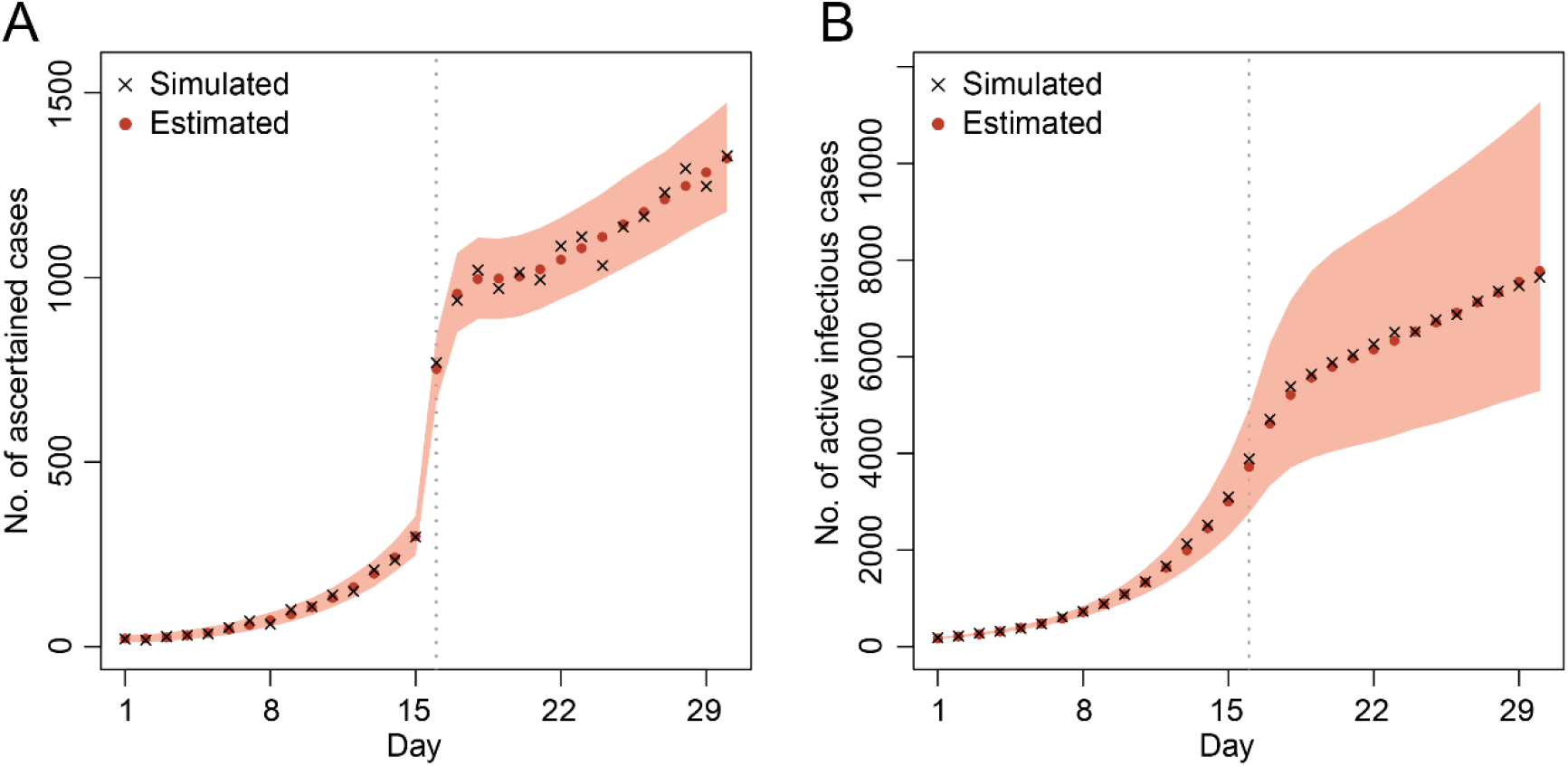
One simulated dataset with two periods. We estimated *b*_1_, *b*_2_, *r*_1_, and *r*_2_ when the other parameters were specified to their true values. (A) Daily incidences. (B) Number of active infectious cases per day, including both ascertained and unascertained cases. The shaded areas indicate 95% CrIs of the estimated values.

**Extended Data Fig. 2.**
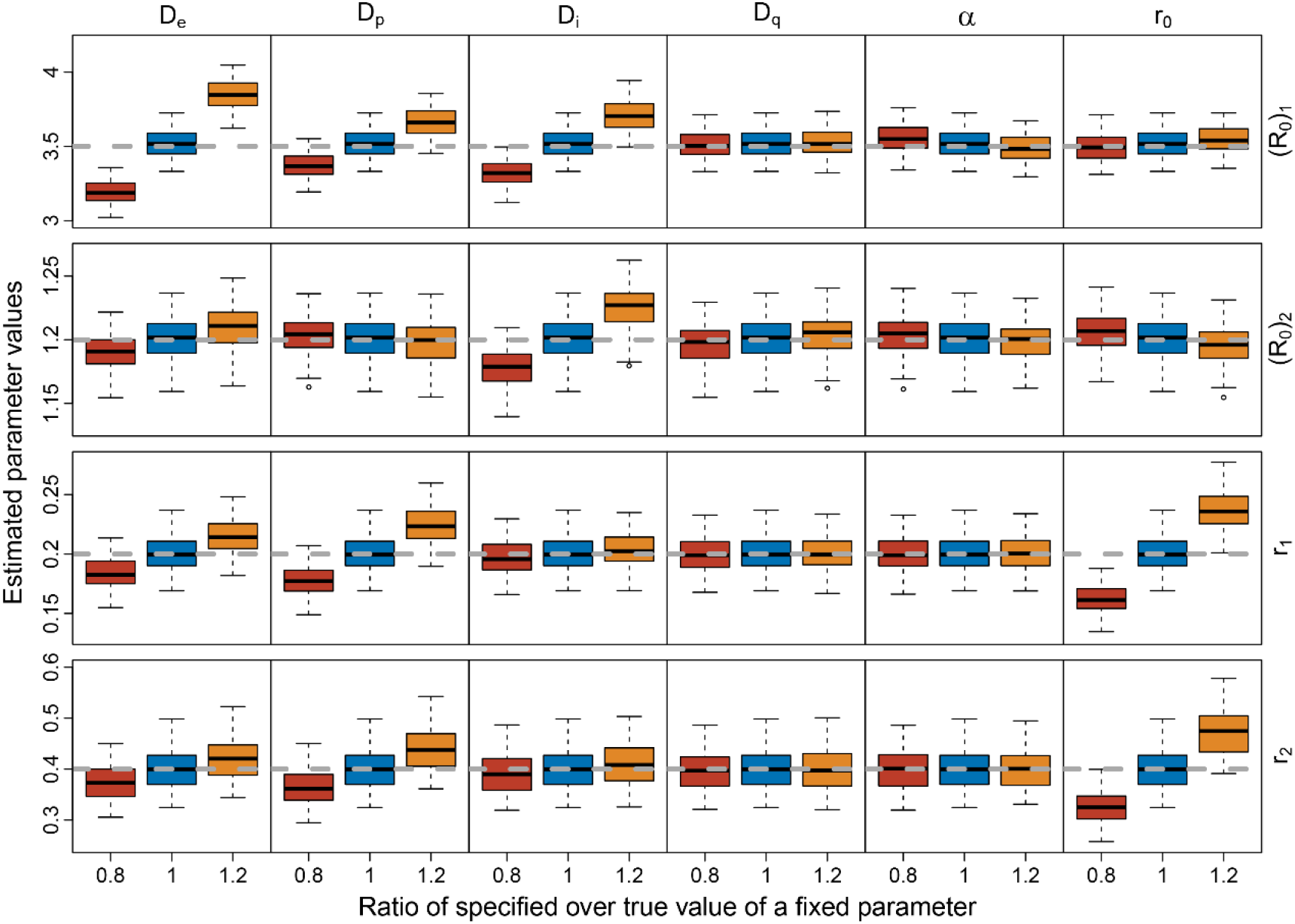
Parameter estimation on simulated epidemic curves with two periods. Each row represents an estimated parameter as indicated on the right, including (*R*_0_)_1_, (*R*_0_)_2_, *r*_1_, and *r*_2_. The grey dashed line in each row represents the true value of the parameter to be estimated. Each column represents a specified parameter as indicated on the top, including *D_e_*, *D_p_*, *D_i_*, *D_q_*, *α*, and *r*_0_, which we specified by the true values or 20% lower or higher than the true values. Each box represents estimates from 100 replicates.

**Extended Data Fig. 3.**
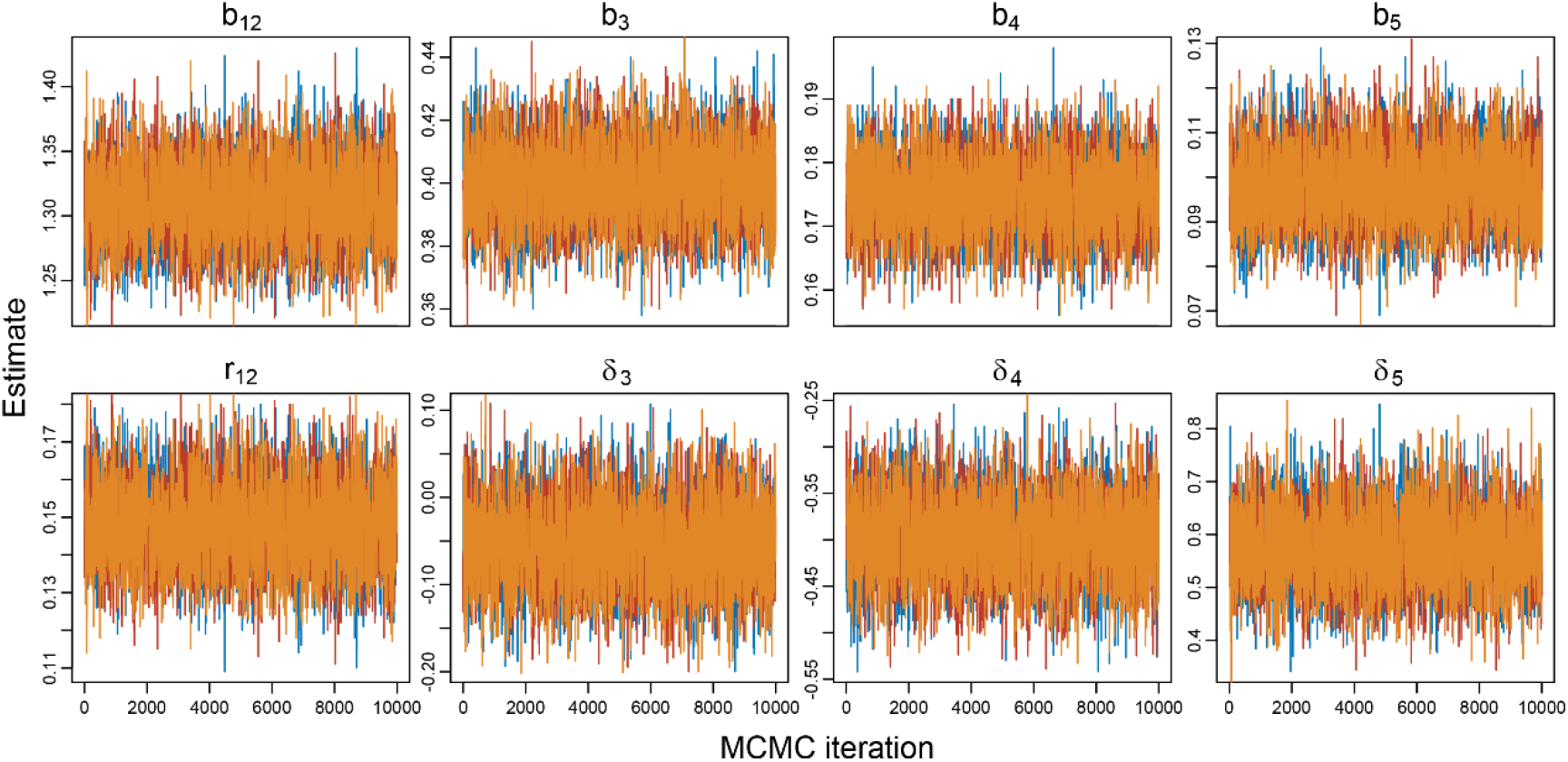
Trace plots of MCMC for the main analysis of real data. Each panel represents the trajectory of 10,000 sampled values for a parameter indicated on the top of the panel. We generated three Markov chains with different initial values, which were colored by orange, red, and blue. The Gelman-Rubin diagnostic was 1.00, indicating convergence of MCMC.

**Extended Data Fig. 4.**
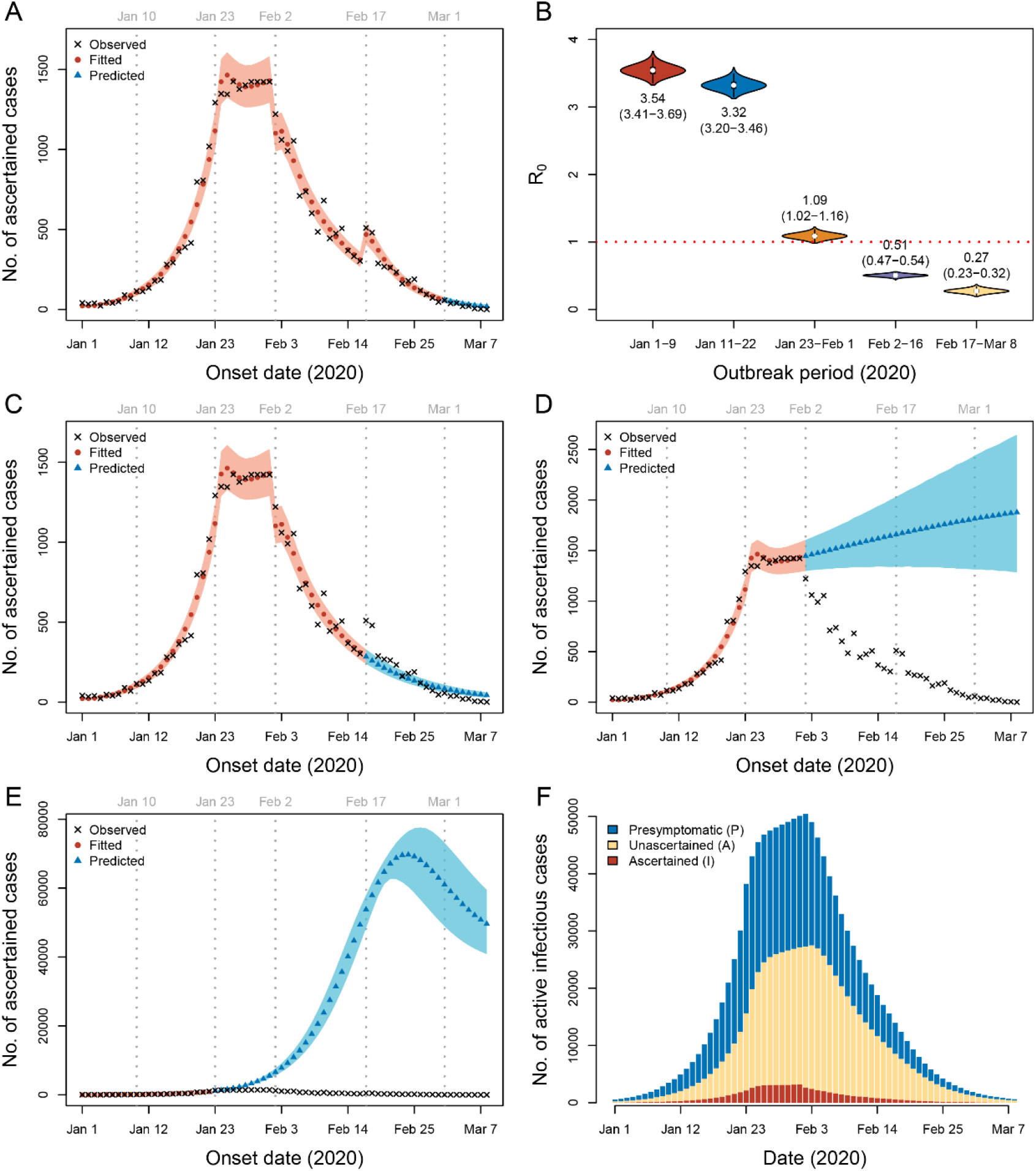
Sensitivity analysis by adjusting the daily incidences from January 29 to February 1 to their average (sensitivity analysis S1) Parameters were estimated by fitting data from January 1 to February 29. (A) Prediction using parameters from period 5 (February 17-29). (B) Estimated R_0_ for each period. The mean and 95% CrI (in parentheses) are labeled below or above the violin plots. (C) Prediction using parameters from period 4 (February 2-16). (D) Prediction using parameters from period 3 (January 23-February 1). (E) Prediction using parameters from period 2 (January 10-22). The shaded areas in (A, C, D and E) are 95% CrI. (F) Estimated number of active infectious cases in Wuhan from January 1 to March 8.

**Extended Data Fig. 5.**
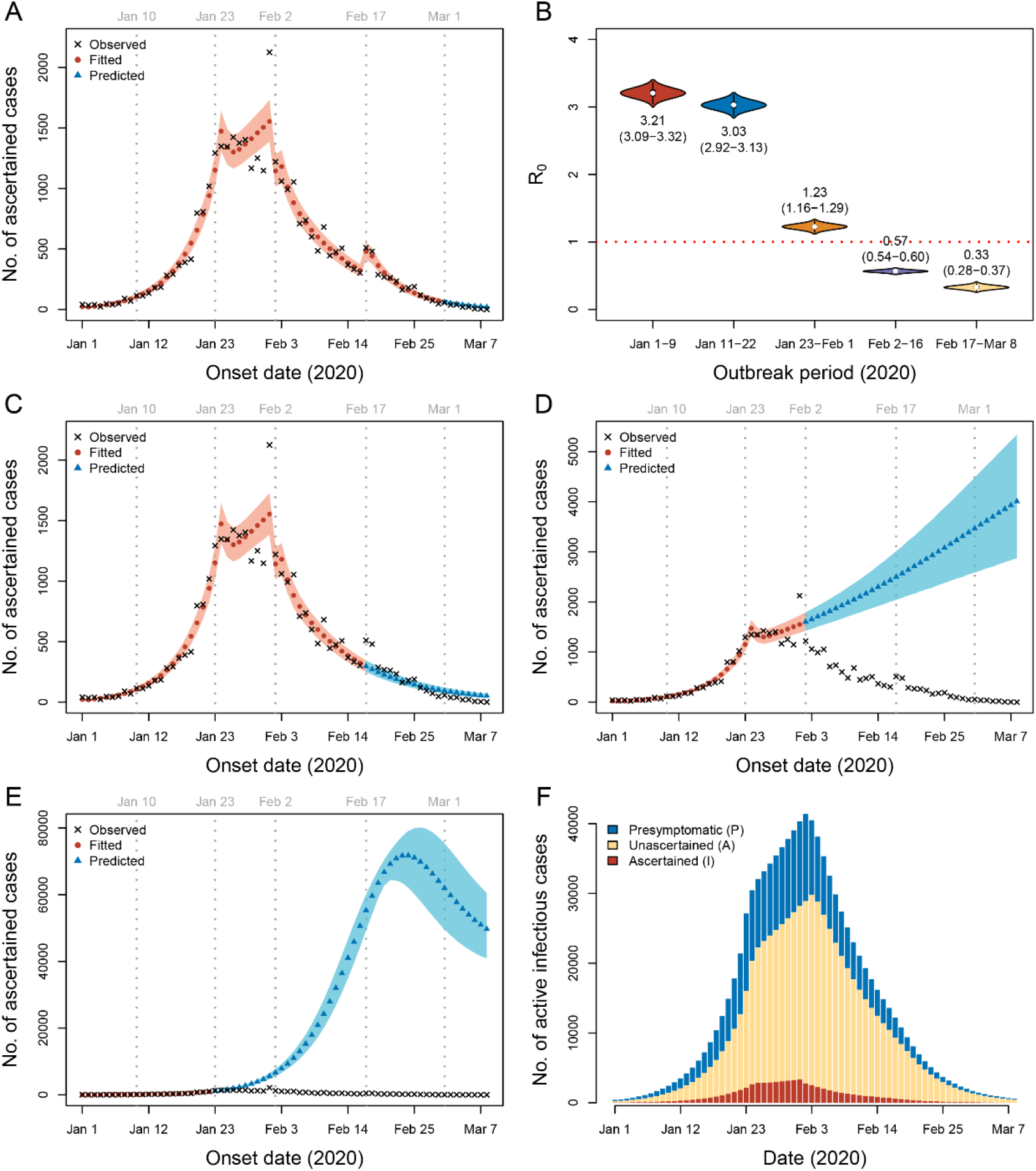
Sensitivity analysis assuming an incubation period of 4.1 days and a presymptomatic infectious period of 1.1 days (sensitivity analysis S2) Parameters were estimated by fitting data from January 1 to February 29. (A) Prediction using parameters from period 5 (February 17-29). (B) Estimated R_0_ for each period. The mean and 95% CrI (in parentheses) are labeled below or above the violin plots. (C) Prediction using parameters from period 4 (February 2-16). (D) Prediction using parameters from period 3 (January 23-February 1). (E) Prediction using parameters from period 2 (January 10-22). The shaded areas in (A, C, D and E) are 95% CrI. (F) Estimated number of active infectious cases in Wuhan from January 1 to March 8.

**Extended Data Fig. 6.**
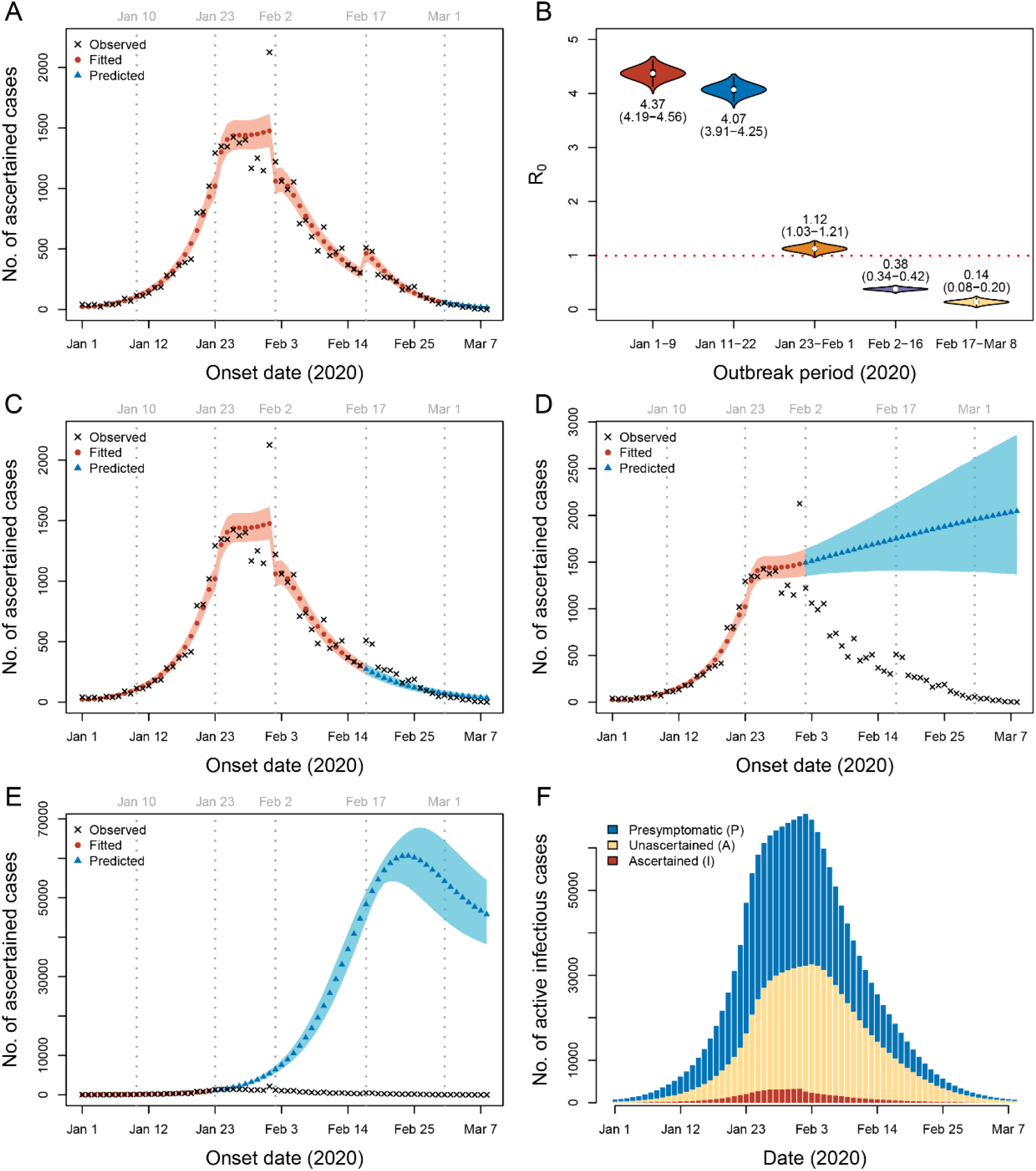
Sensitivity analysis assuming an incubation period of 7 days and a presymptomatic infectious period of 3 days (sensitivity analysis S3) Parameters were estimated by fitting data from January 1 to February 29. (A) Prediction using parameters from period 5 (February 17-29). (B) Estimated R_0_ for each period. The mean and 95% CrI (in parentheses) are labeled below or above the violin plots. (C) Prediction using parameters from period 4 (February 2-16). (D) Prediction using parameters from period 3 (January 23-February 1). (E) Prediction using parameters from period 2 (January 10-22). The shaded areas in (A, C, D and E) are 95% CrI. (F) Estimated number of active infectious cases in Wuhan from January 1 to March 8.

**Extended Data Fig. 7.**
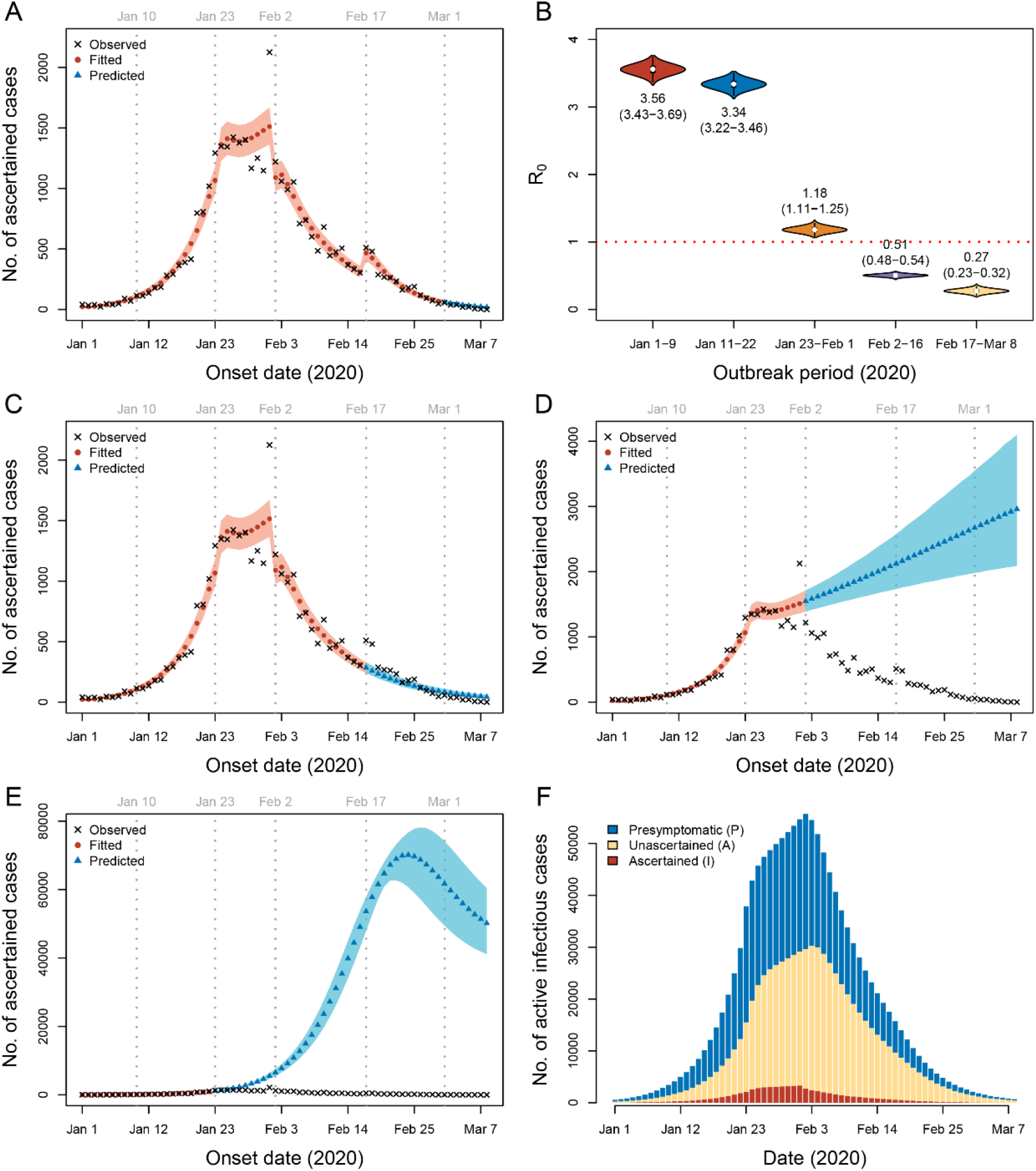
Sensitivity analysis assuming the transmissibility of the presymptomatic and unascertained cases is 0.46 of the ascertained cases (sensitivity analysis S4) Parameters were estimated by fitting data from January 1 to February 29. (A) Prediction using parameters from period 5 (February 17-29). (B) Estimated R_0_ for each period. The mean and 95% CrI (in parentheses) are labeled below or above the violin plots. (C) Prediction using parameters from period 4 (February 2-16). (D) Prediction using parameters from period 3 (January 23-February 1). (E) Prediction using parameters from period 2 (January 10-22). The shaded areas in (A, C, D and E) are 95% CrI. (F) Estimated number of active infectious cases in Wuhan from January 1 to March 8.

**Extended Data Fig. 8.**
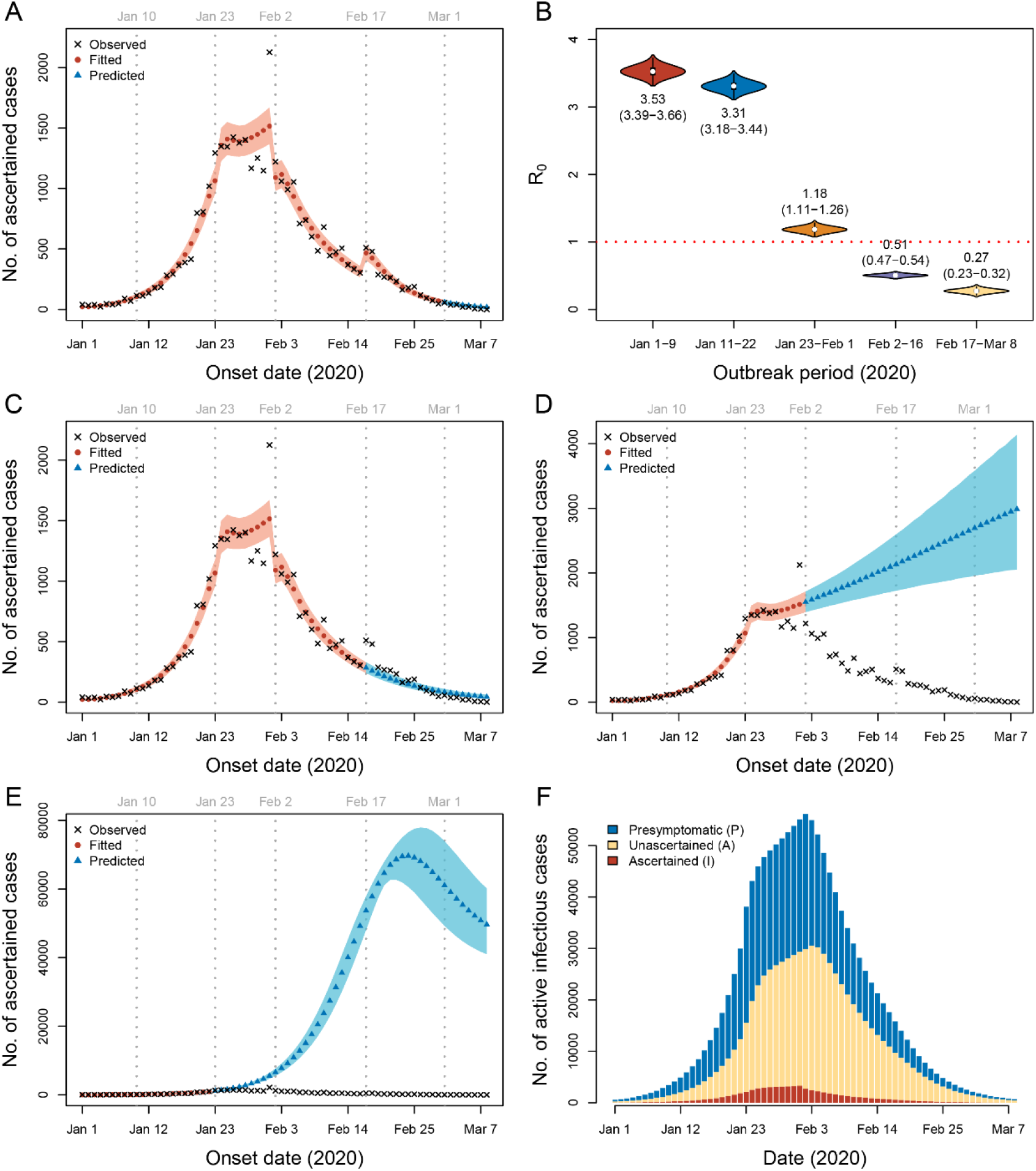
Sensitivity analysis assuming the transmissibility of the presymptomatic and unascertained cases is 0.62 of the ascertained cases (sensitivity analysis S5) Parameters were estimated by fitting data from January 1 to February 29. (A) Prediction using parameters from period 5 (February 17-29). (B) Estimated R_0_ for each period. The mean and 95% CrI (in parentheses) are labeled below or above the violin plots. (C) Prediction using parameters from period 4 (February 2-16). (D) Prediction using parameters from period 3 (January 23-February 1). (E) Prediction using parameters from period 2 (January 10-22). The shaded areas in (A, C, D and E) are 95% CrI. (F) Estimated number of active infectious cases in Wuhan from January 1 to March 8.

**Extended Data Fig. 9.**
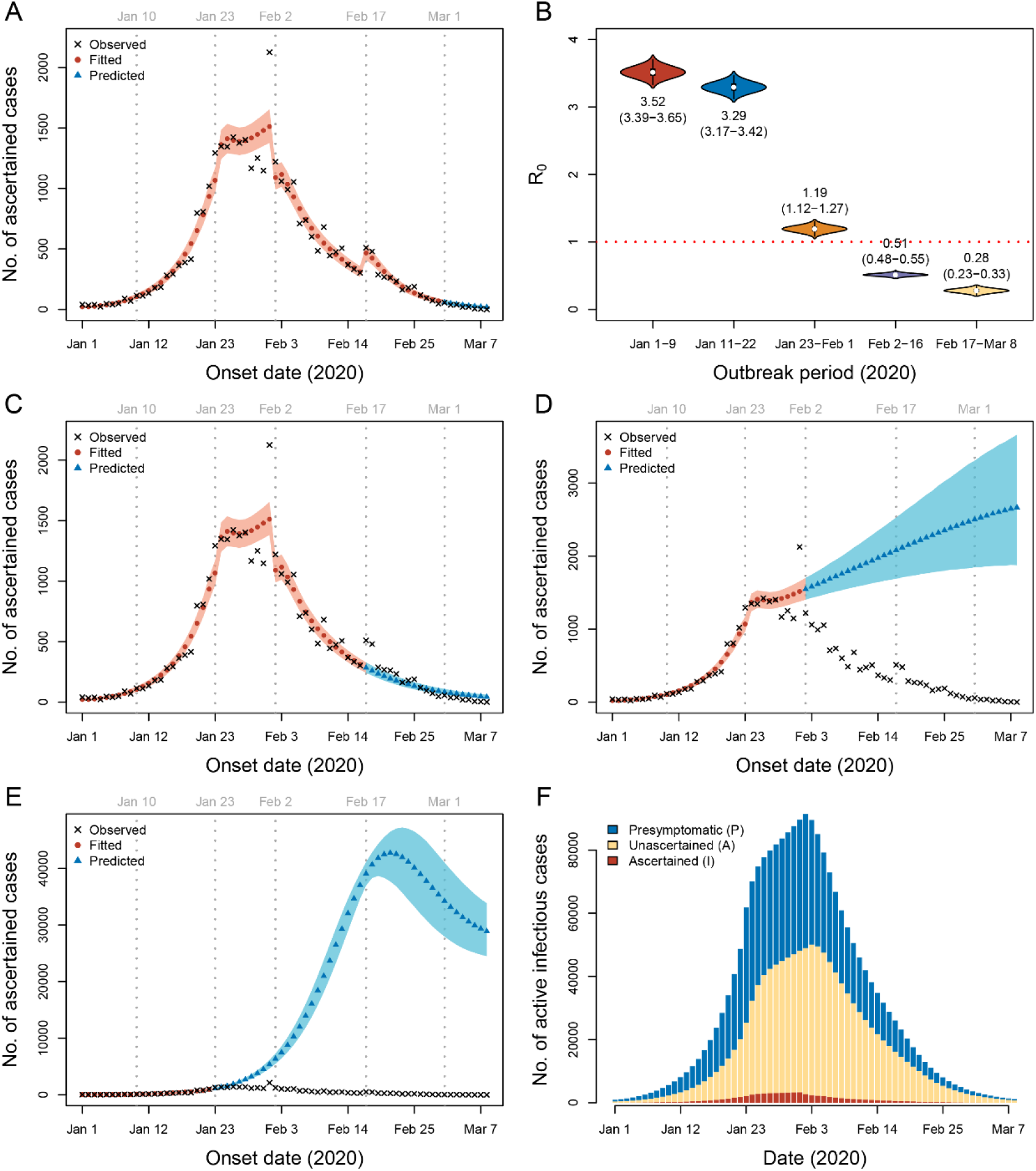
Sensitivity analysis assuming the initial ascertainment rate is *r*_0_ = 0.14 (sensitivity analysis S6) Parameters were estimated by fitting data from January 1 to February 29. (A) Prediction using parameters from period 5 (February 17-29). (B) Estimated R_0_ for each period. The mean and 95% CrI (in parentheses) are labeled below or above the violin plots. (C) Prediction using parameters from period 4 (February 2-16). (D) Prediction using parameters from period 3 (January 23-February 1). (E) Prediction using parameters from period 2 (January 10-22). The shaded areas in (A, C, D and E) are 95% CrI. (F) Estimated number of active infectious cases in Wuhan from January 1 to March 8.

**Extended Data Fig. 10.**
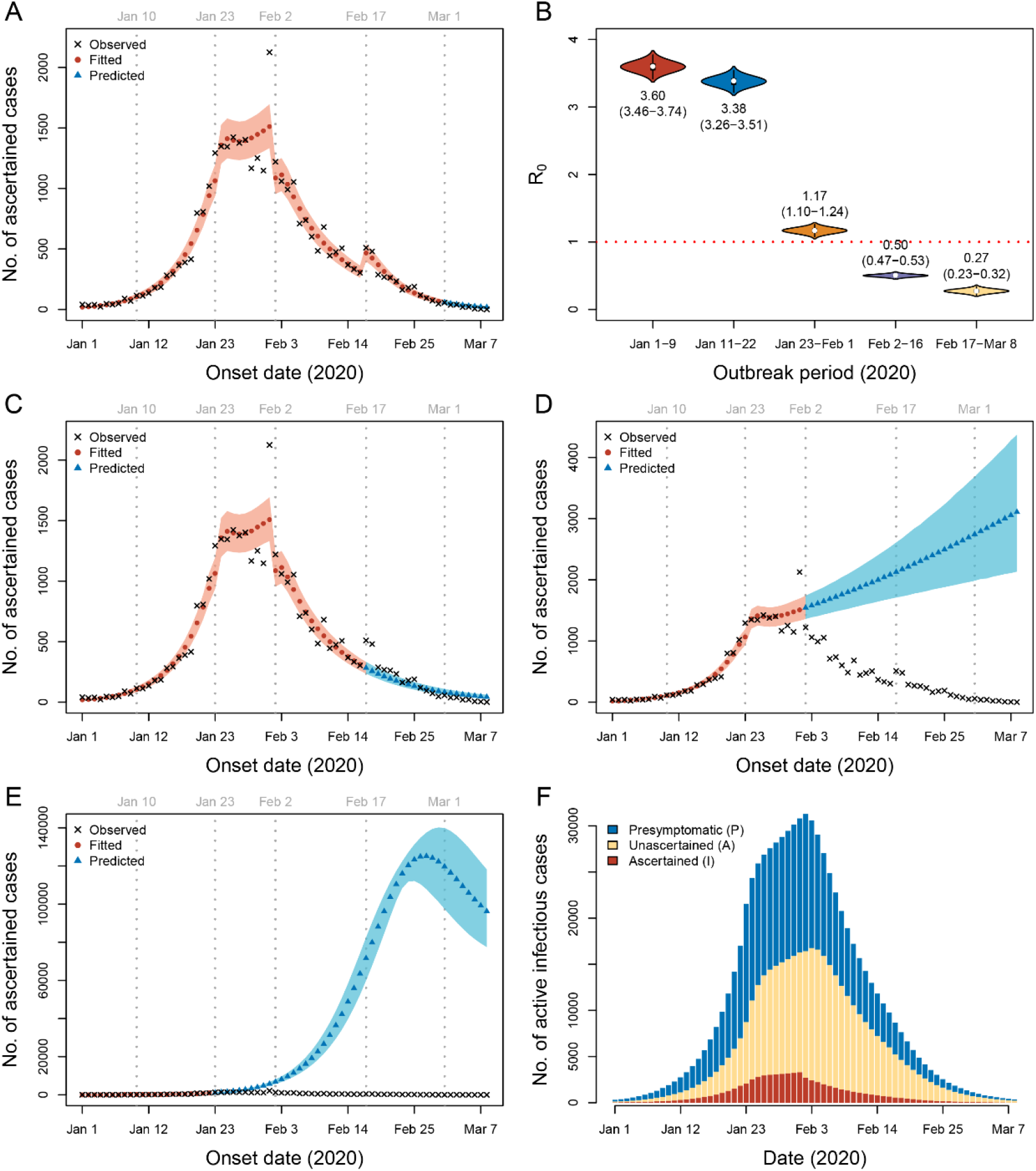
Sensitivity analysis assuming the initial ascertainment rate is *r*_0_ = 0.42 (sensitivity analysis S7) Parameters were estimated by fitting data from January 1 to February 29. (A) Prediction using parameters from period 5 (February 17-29). (B) Estimated R_0_ for each period. The mean and 95% CrI (in parentheses) are labeled below or above the violin plots. (C) Prediction using parameters from period 4 (February 2-16). (D) Prediction using parameters from period 3 (January 23-February 1). (E) Prediction using parameters from period 2 (January 10-22). The shaded areas in (A, C, D and E) are 95% CrI. (F) Estimated number of active infectious cases in Wuhan from January 1 to March 8.

**Extended Data Fig. 11.**
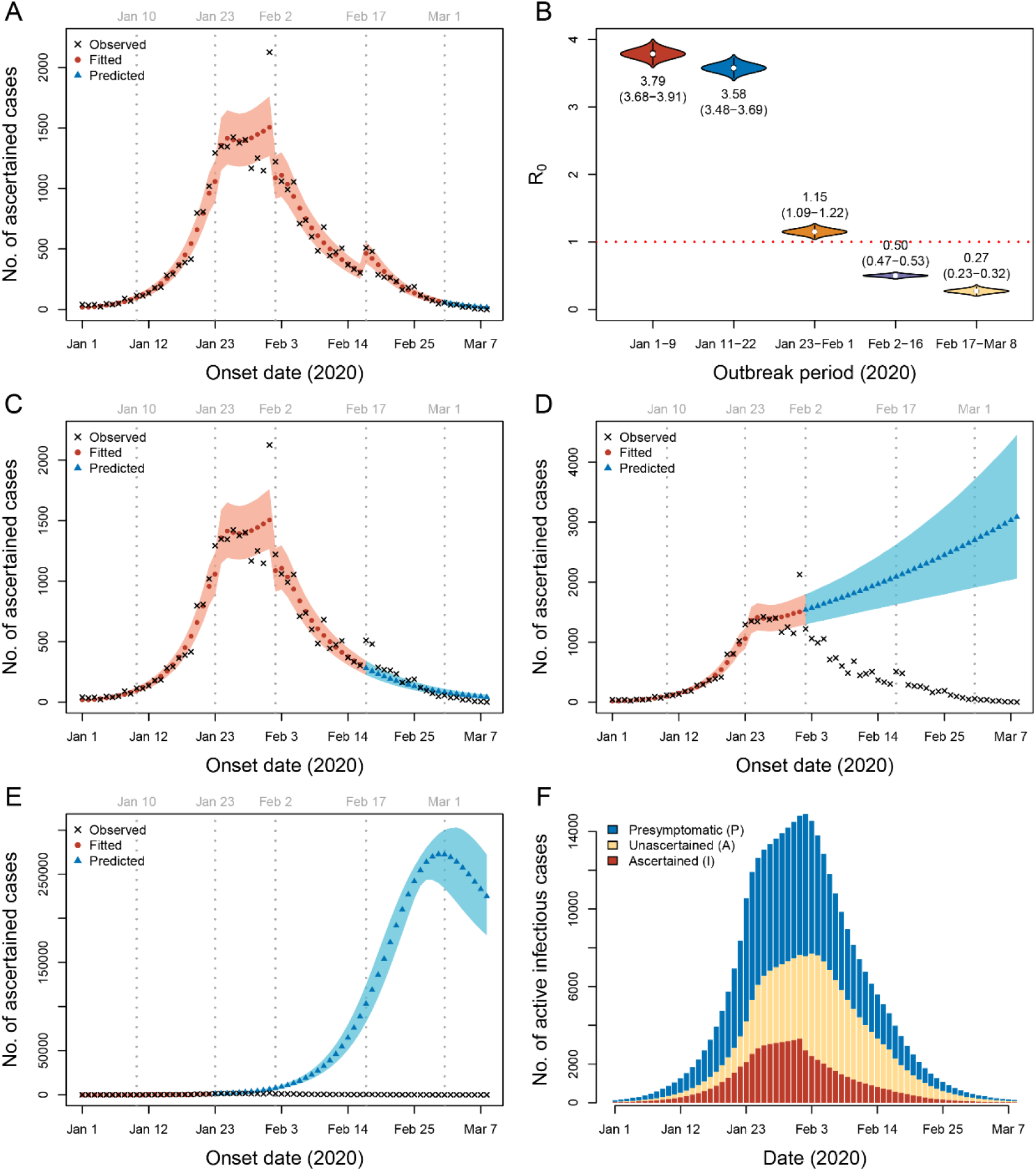
Sensitivity analysis assuming the initial ascertainment rate is *r*_0_ = 1 (sensitivity analysis S8) Parameters were estimated by fitting data from January 1 to February 29. (A) Prediction using parameters from period 5 (February 17-29). (B) Estimated R_0_ for each period. The mean and 95% CrI (in parentheses) are labeled below or above the violin plots. (C) Prediction using parameters from period 4 (February 2-16). (D) Prediction using parameters from period 3 (January 23-February 1). (E) Prediction using parameters from period 2 (January 10-22). The shaded areas in (A, C, D and E) are 95% CrI. (F) Estimated number of active infectious cases in Wuhan from January 1 to March 8.

**Extended Data Fig. 12.**
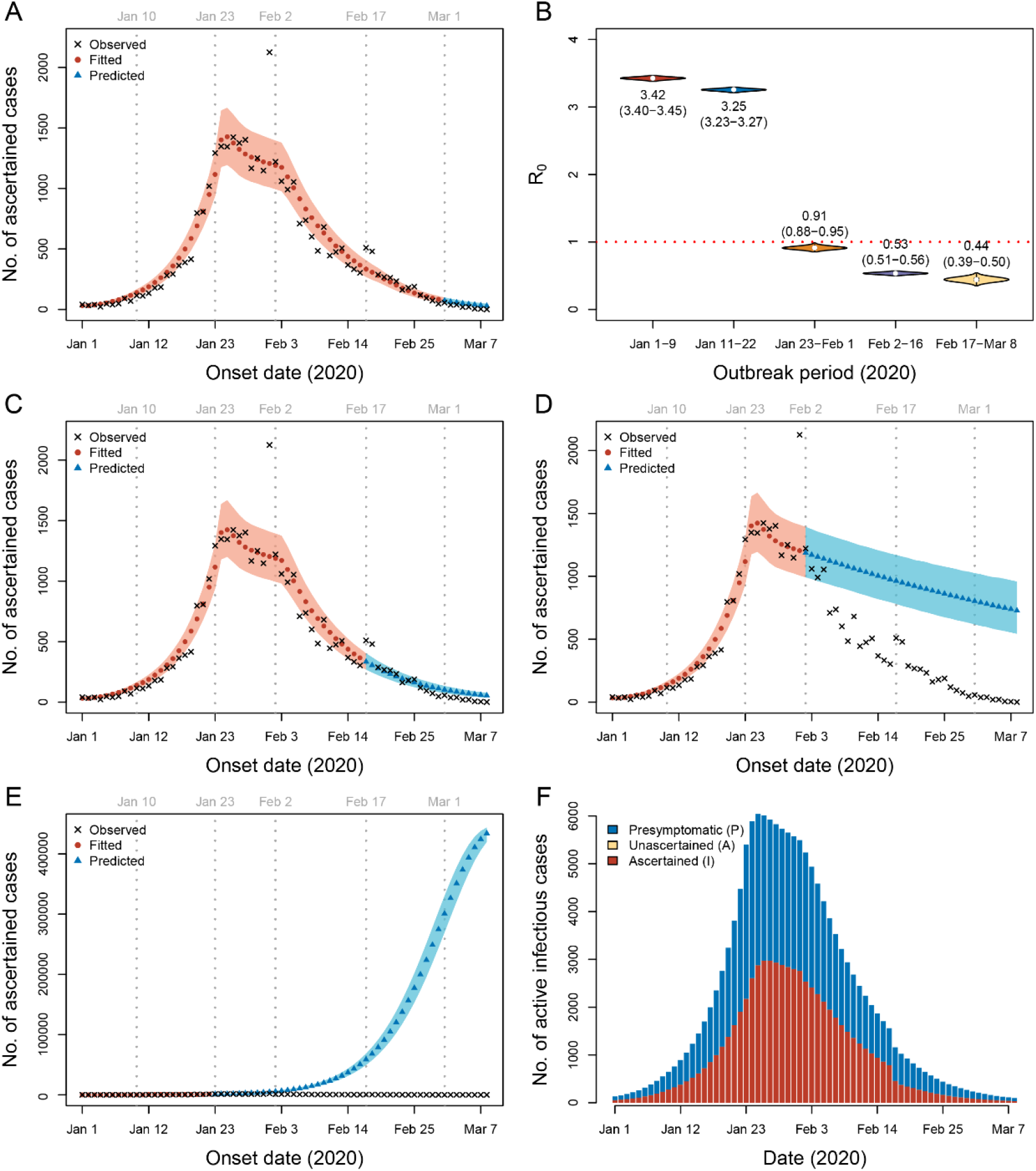
Sensitivity analysis assuming complete ascertainment at any time (sensitivity analysis S9) Parameters were estimated by fitting data from January 1 to February 29. Compared to the full model, this simplified model fit the data significantly worse (likelihood ratio test, 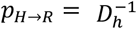, *p* = 0). (A) Prediction using parameters from period 5 (February 17-29). (B) Estimated R_0_ for each period. The mean and 95% CrI (in parentheses) are labeled below or above the violin plots. (C) Prediction using parameters from period 4 (February 2-16). (D) Prediction using parameters from period 3 (January 23-February 1). (E) Prediction using parameters from period 2 (January 10-22). The shaded areas in (A, C, D and E) are 95% CrI. (F) Estimated number of active infectious cases in Wuhan from January 1 to March 8.

**Extended Data Fig. 13.**
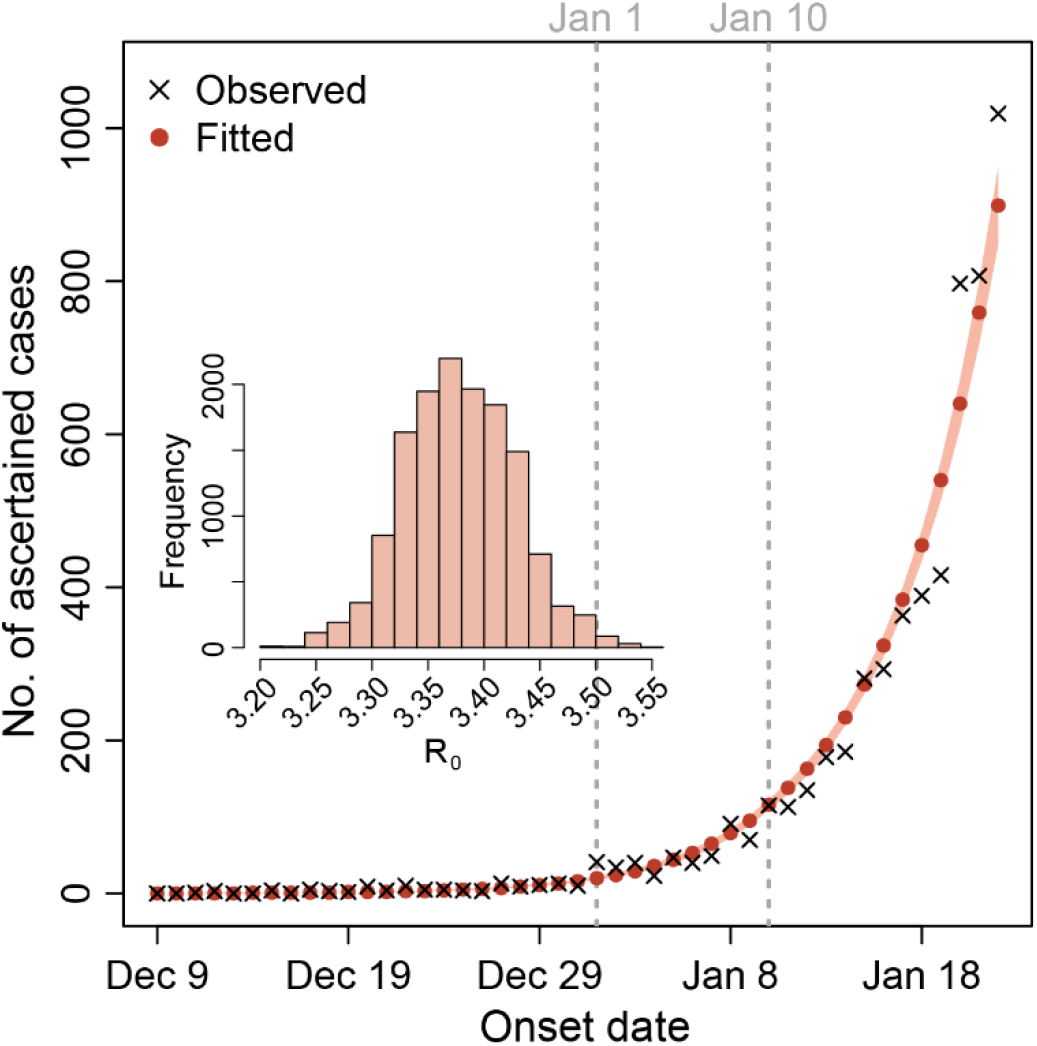
Estimation of R_0_ using daily incidence data starting from December 9. Following the main analysis, we assumed *r*_0_ = 0.23 and set *I*(0) = 1, *A*(0) = 3, *E*(0) = 17 and *P*(0) = *H*(0) = *R*(0) = 0 accordingly. We assumed transmission rate *b*, ascertainment rate *r*, and duration from illness onset to hospitalization *D_q_* (set to 21 days) were the same until January 22, 2020. All the other settings were the same as in the main analysis. The shaded area in the plot indicates 95% CrIs estimated by the deterministic model with 10,000 sets of parameter values sampled from MCMC. Unlike other analyses, we did not construct 95% CrIs by stochastic simulations, because stochastic fluctuations at the early days would have extremely large impacts due to low counts, leading to unreasonable CrIs. The inserted histogram shows the distribution of the estimated R_0_ from December 9, 2019 to January 22, 2020, for which the mean estimate was 3.38 (95% CrI: 3.28-3.48).

